# Application of Digital Twin and Heuristic Computer Reasoning to Workflow Management: Gastroenterology Outpatient Centers Study

**DOI:** 10.1101/2022.03.22.22272507

**Authors:** Marc Garbey, Guillaume Joerger, Shannon Furr

## Abstract

The workflow in a large medical procedural suite is characterized by high variability of input and suboptimum throughput. Today, Electronic Health Record systems do not address the problem of workflow efficiency: there is still high frustration from medical staff who lack real-time awareness and need to act in response of events based on their personal experiences rather than anticipating. In a medical procedural suite, there are many nonlinear coupling mechanisms between individual tasks that could wrong and therefore is difficult for any individual to control the workflow in real-time or optimize it in the long run. We propose a system approach by creating a digital twin of the procedural suite that assimilates Electronic Health Record data and supports the process of making rational, data-driven, decisions to optimize the workflow on a continuous basis. In this paper, we focus on long term improvements of gastroenterology outpatient centers as a prototype example and use six months of data acquisition in two different clinical sites to validate the artificial intelligence algorithms.

## 1. Introduction

This paper addresses the challenge of optimizing the quality and throughput of the workflow in a complex environment, such as surgical suites.

As of today, medical staff working daily in a surgical suite are still struggling with obvious questions, such as: Why can’t we anticipate problems? Why is our utilization low? How can we effectively treat more patients? Our staff is stressed, they need more control, how do they get it? Why is the preparation of patients so slow? Why don’t we ever start on time? etc. [chapter 31 in [1]].

There is a very large number of publications on workflow optimization ranging from purely descriptive statistical methods to manufacturing industry process methods [2,3,4]. Recently, machine learning techniques, deep learning in particular, have gained traction [5].

However, there are few fundamental difficulties in the problem to be addressed that have limited the success of all these methods:

i. The dataset comes from the Electronic Health Record (EHR), and the quality and accuracy of the data might be highly questionable [7,8,9].
ii. There is not a consistent workflow routine at the daily/monthly time scale in healthcare: conditions keep changing because of the population of patient variability, large turnover of staff, and other external events (COVID-19, reimbursement drivers, and evolving tools/processes). Consequently, the sample dataset one can work with for computational modeling is small by nature [10].
iii. Every clinic or hospital might be different depending on staff, management models, and facility layouts: there might be no reliable global rules that apply nor one solution that fits all [11].

From a system science point of view, surgical flow in a large facility is a complex nonlinear system with multiple decision trees, multi-factorial processes, and interactions, which is further intensified and exacerbated by human factors between all involved “players.” Manifestly, it is only long-term experiences in a specific surgical facility that helps a good manager of the floor to fully appreciate and anticipate problems in order to formulate and execute the right decisions for the management of a facility. Each facility has its own pace: one operating mode is usually not applicable to another facility as human behaviors, environments, and modes of operation are highly location specific [11]. An essential feature of our solution process is the construction of the so-called digital twin of the clinic’s workflow. A digital twin is “a virtual representation that serves as the real-time digital counterpart of a physical object or process” and was first introduced for manufacturing processes by NASA to improve physical model simulations of spacecraft in 2010 [12].

The digital twin is fit to the clinic’s workflow with respect to the probability distribution of the main metrics (throughput and safety) that correspond to the focus of the management team. Upon it, we built a heuristic computational reasoning set of algorithms that act as a “What-if questioning virtual machine.” The resulting system is curious of what may go wrong or might be better based on previous observations and benchmarking. It continuously runs a multitude of scenarios using the digital twin in order to optimize the workflow [13,14,15]. Now, managers and staff may combine their experiences with the recommendations provided by the heuristic computer reasoning algorithm exercising the digital twin analysis to make informed, data-driven decisions that have better chances of success.

The solution described in this paper concentrates on gastrointestinal (GI) procedural suites in outpatient centers as our prototype example [16]. This approach may apply to many categories of specialized procedural suites, from catheterization laboratory to ophthalmology, from orthopedic centers to bariatric centers, that share the same type of organized workflow with high volume routine procedures.

GI’s diagnostic laboratories are less complex than a general surgery floor that includes dozens of different types of procedures and has corresponding workflows with multiple paths involving medical imaging and biological labs. GI outpatient centers deliver two main categories of quick procedures: colonoscopy and EsophagoGastroDuodenoscopy (EGD). The patient workflow is a one-way linear graph, exhibiting standard workflow steps from registration to discharge and “rendezvous” points including the cycle of scope management - see Figure 3. Thus, number of unknowns and independent variables in a digital model of an outpatient GI center are lower compared to a general surgical suite; however, (i) to (iii) still hold true in GI outpatient centers with multiple procedure rooms. For example, Figure 1 and Figure 2 show the typical rate of error of EHR timestamps in a GI center and how it can relate to human fatigue throughout the day. Consequently, the problem of GI workflow optimization remains challenging with accumulating gain or loss at the minute scale.

**Figure 1:**
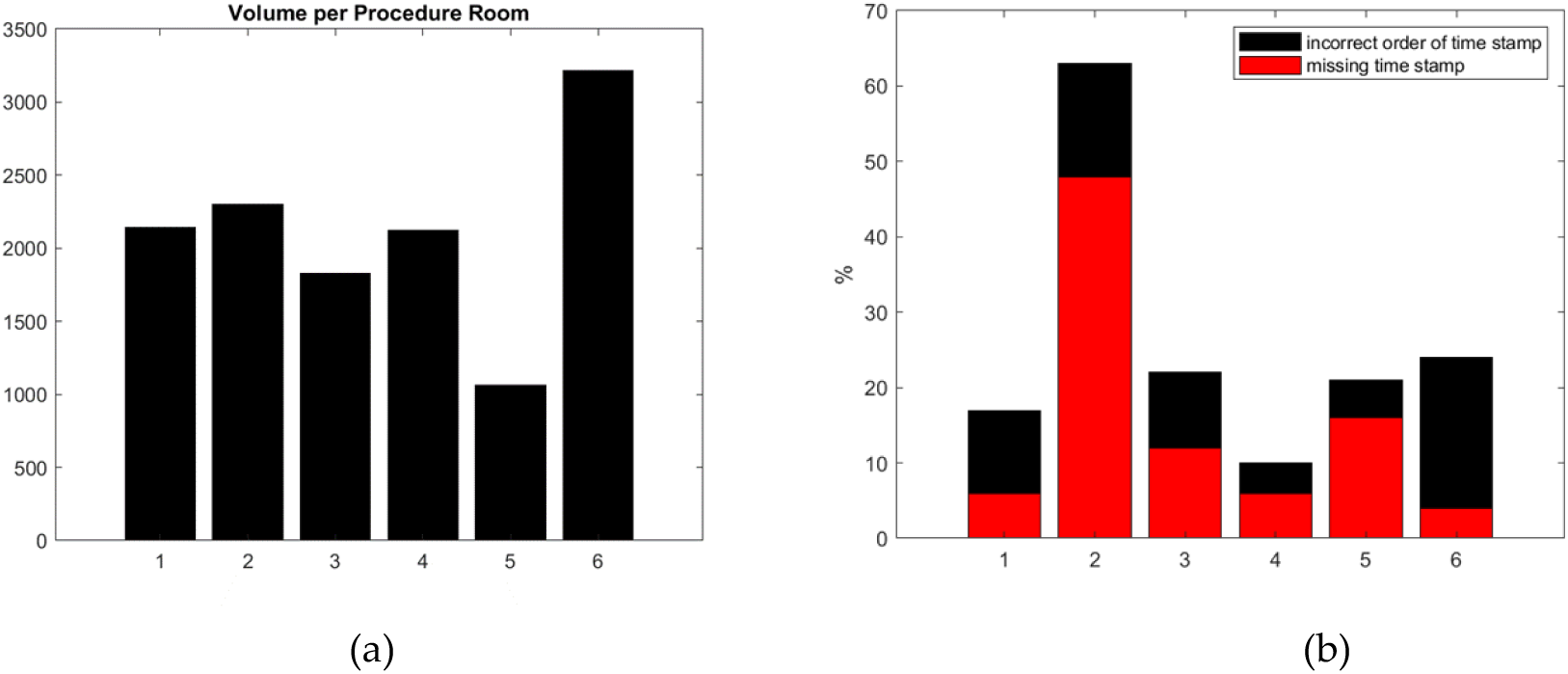
(a) Estimated volume of patients per procedure room per year in each outpatient center (rate of error is not correlated to volume). (b) Basic error detection rate based on missing entries or incoherent order of timestamps at each outpatient center. Other types of errors are possible but not counted here.

**Figure 2:**
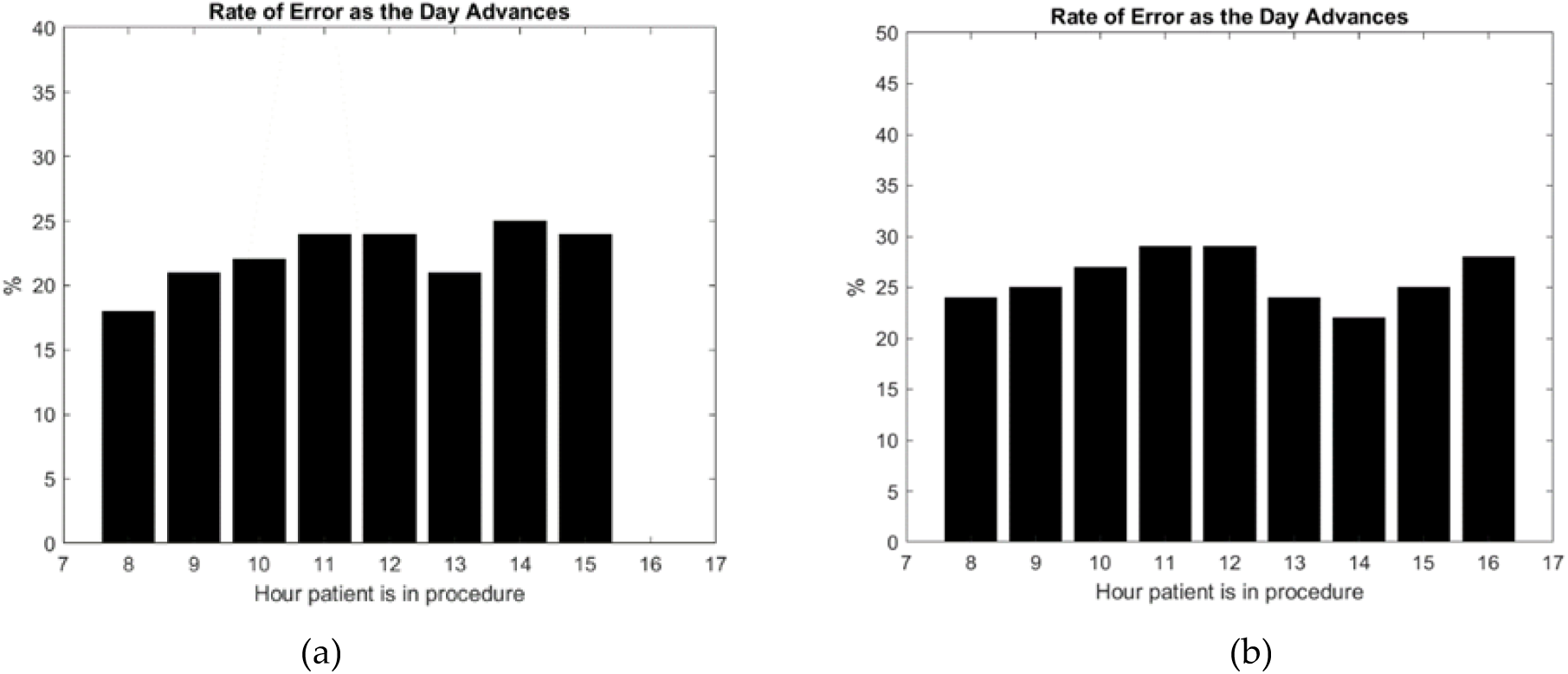
An example of the correlation found between clinical working hours and rate of error occurrence in two outpatient centers (a) and (b). The percentage of error represents the number of errors per hour weighted by the number of patients per hour in procedure room. The error rate increases during the morning with fatigue accumulation, marked with a pause at lunch time, and then increases again until the end of the clinical day.

**Figure 3:**
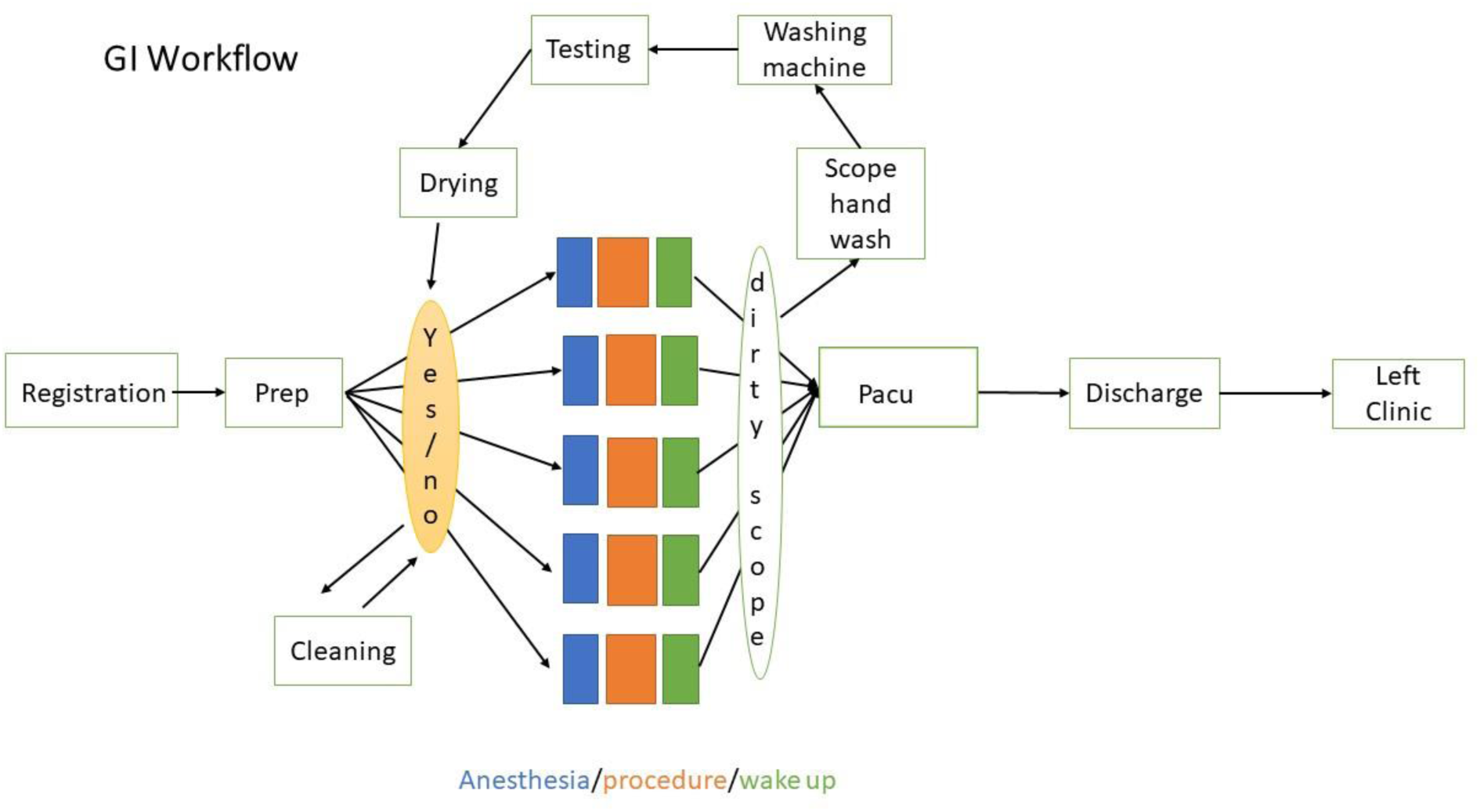
Workflow of a Gastroenterology Clinic.

GI diagnostic procedures are usually short, about 20 minutes for a colonoscopy procedure, 10 minutes for an EGD. Typical billing for these is about $1K if one weights Medicare volume with private insurance volume reimbursement. According to a John Hopkins report on infection rate after procedures, colonoscopy (respectively EGD) represents about 15 million (respectively 7 million) procedures per year in the USA in 2018. Advanced GI procedures involving therapeutic treatments can be longer and are less predictable: an Endoscopic Retrograde CholangioPancreatography (ERCP) might take between 30 minutes to 2 hours depending on the complexity, but the reimbursement is much higher than general GI procedures. The number of advanced procedures will continue to increase because the number of traditionally invasive procedures is decreasing to allow for minimally invasive procedures.

Hospitals handle GI diagnostics and GI procedures that are more complex, and declare difficult patient conditions relatively more than outpatient centers. This paper is focusing on outpatient centers, and we will present the digital twin of a hospital’s GI lab in a future publication [17,18,19,20]. The cost to run GI procedures has been increasing faster than reimbursement. Consequently, GI labs are interested in restoring their previous financial position by increasing throughput and/or lowering costs. Hence, they are not only interested in revenue. Because of the high turnover of nurses and providers, GI labs want to provide good working conditions within a well-run infrastructure to improve staff retention. GI labs may compete with each other to attract new patients. Patient satisfaction as well as quality of care are key criteria for patient retention and reimbursement rates. Patients expect to experience an efficient and caring workflow as well, so this also holds the potential of lowering the costs to treat patients and more robust finances for the clinic.

In the next section, we will describe our general methodology to address the problem of workflow optimization in specialized medical procedural suites, starting from the digital twin concept with agent-based modeling [17,21,22].

## 2. Materials and Methods

We are going to first introduce the digital twin of the clinic’s workflow, which is the main computational engine of the workflow optimization technique used to support workflow management.

### 2.1 Digital Twin

A digital twin is a virtual representation that serves as the real-time digital counterpart of a physical object or process. The digital twin may combine multiple models, may require the integration of heterogonous data sets, and may take into account human factors. There are similarities in the limitations of digital twin and computational model in general: both can be very inaccurate unless proved to be valid.

One can refer to the quotation of George E.P. Box: “Essentially all models are wrong, but some are useful.” First, we design our digital twin to deliver adequate accuracy for a specific purpose. Our purpose for the digital twin of a GI lab is to support the staff in anticipating workflow bottlenecks in their daily practice, assisting in root cause analysis of problems, guiding the manager to make strategic changes, and/or recruiting to improve workflow according to an objective function. Such objectives, without limitation, can be:

i. increasing the number of patients processed during the clinic day,
ii. improving on-time workflow steps related to late starts, turnover, or overtime,
iii. improving quality by allowing each provider to spend enough time with patients.

Every clinic may attribute a different weight to each of these three goals. Second, we refer to the Ockham’s razor principle as reported in Albert Einstein’s famous quotation: “Everything [in a model] should be made as simple as possible, but not simpler.” A digital twin has intrinsically higher complexity than a model but should be made as simple as possible while still being “useful” to reach, in our specific GI application this is our workflow improvement objective. The more complex the digital twin is, the more input data it may need and the less these data might be accurate. A digital twin as complex as the clinical process would be and as unmanageable as the clinic process itself is.

Third, the quality of the model depends on the quality of the input data as stated in the popular quote “garbage in, garbage out.” This is very relevant to a digital twin that depends on a disparate input information, as it is intended to cover all aspects of the physical object or process. For our specific application, we need timestamps of workflow usually entered manually in the EHR system, scheduling information provided by a third-party software, team composition, some characteristics of staff behavior and performance. We use a simple mathematical framework to model human behavior affecting the multiple steps of the workflow from registration to discharge based on descriptive statistics of individual performances.

To build the digital twin of the GI lab, we start from a graphical representation of the workflow – see Figure 3. Each node corresponds to a step of the process. The arrows of the directed graph give the order of tasks in which they must be achieved. The patient arrives at registration and, once their information is entered into the EHR system, a nurse is ready to bring the patient to the pre-op area. Once the patient has been prepared for the procedure, the patient moves to the procedure room assigned to their provider. Inside the procedure room, the process generally starts with anesthesia, goes through the procedure itself and ends with the patient waking up (end of anesthesia). But different GI labs may have different practices in anesthesia with respect to possible start, respectively ending of the anesthesia work, in preparation unit (Prep), respectively Post Anesthesia Care Unit (PACU). If there is room available in PACU and a nurse ready to assist with recovery, the patient moves to PACU. They are finally discharged and leave the clinic if their ride is ready for patient pick up.

Every step is conditioned on the premise of availability of specific resources: (1) that the required staff are available to do the task and (2) the equipment is prepped and ready for the procedure. For example, the flexible scope processing, which is central to GI labs, has its own cycle and needs to be coordinated with the procedure steps as illustrated in the upper loop of Figure 3.

The design architecture of the digital twin is such that it can take the current global state of the clinic, including room state and key locations, and predicts what would be its state at a point later in time.

The digital twin is not the end-result by itself; it needs to be set in a framework that can make it useful. Figure 4 describes how the digital twin’s computational engine is embedded into the architecture of a heuristic computer reasoning system used to support the workflow management goal.

**Figure 4:**
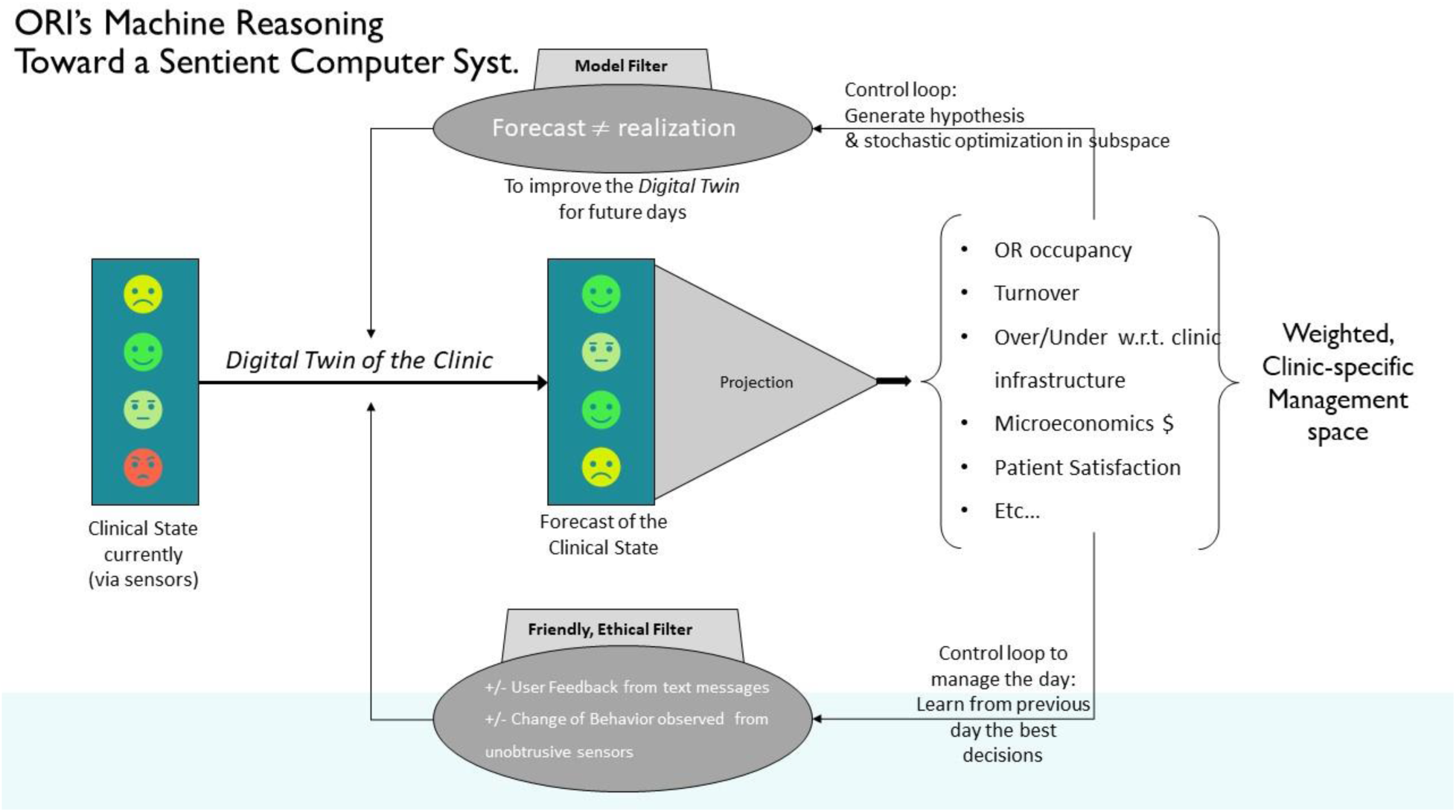
Architecture of our HERLAD System

To define the objective function of the digital twin, we are using a projection operator that computes the various metrics expressing operable efficiencies of the workflow. As often as needed, an optimization loop (see upper loop in Figure 4) fits the digital twin’s predictions to the observation, adjusting the free parameters of the model. This process is stochastic (randomly determined) since the prediction itself has randomness and the digital twin needs to be ran multiple times to generate statistics on real versus potential output. We use a combination of a genetic algorithm and a swarm algorithm to accelerate the convergence of the optimization algorithm [23]. Overall metrics of efficiency and safety are weighted according to the institution’s needs, under certain ethical constraints. For example, financial profitability of the GI lab should not be the only objective in the short term, since patient safety and satisfaction must be part of the objectives. In this aspect, the digital twin runs the forward scenarios multiple times, simultaneously adjusting specific control parameters through a multitude of defined ranges, to present to a manager or operator a framework of potential outcomes and endpoints from highest to lowest probability where distinct parameters are highly variable and heavily influenced by human inputs and interactions. Moreover, the system uses both current and historical data (as described above) in order to capture present data, compare current versus historic data and detect trends, when possible, of real-time and possible future workflows. The system uses multiple constraints in the optimization process (see lower loop in Figure 4) to take into account not only movements and activities of each variable (patients treatments, staff, equipment, and space) but also incorporates staff feedback and patient feedback entered through the interface, or changes of behavior observed through the sensors of the system, to serve the two-fold function of (1) enhancing the predictive value of the system while (2) ensuring that the system operates under admissible conditions in terms of ethical standards – each effecting potential outcomes and endpoints.

A detailed understanding of the digital twin requires the construction of a mathematical framework that is provided in the appendix. Next, we will describe the data sets that the digital twin of the clinic’s workflow requires for calibration.

### 2.2 Data Acquisition and Calibration of the Digital Twin

To calibrate the digital twin, we first collect timestamps of the workflow that are entered in the EHR system by medical staff. This is standard practice and does not require additional work.

Table 1 describes this data collection. In addition to this series of 11-to-13 timestamps depending on how bidirectional endoscopy procedures (also called doubles) are entered in the system, we get an ID to track providers and staff and have consistency in our individual behavior model. Our algorithm is HIPAA compliant: the data set has been deidentified; specifically, we do not manipulate any patient protected health information (PHI) and we only receive a case ID to crosscheck data coming from the same procedure.

**Table 1:** information extracted from the EHR

| # | Data Element from EHR | Format |
| --- | --- | --- |
| 1 | Procedure *Scheduled* Date & Time | mm/dd/yyyy hh:mm |
| 2 | Procedure Room Number | Text |
| 3 | Type of Procedure | Text |
| 4 | Patient Name (or Case ID) | Text (*anonymized*) |
| 5 | Provider (or ID) | Text (*anonymized*) |
| 6 | CRNA (or ID) | Text (*anonymized*) |
| 7 | Type of Anesthesia | Text |
| 8 | Time: Admit/Front Desk | hh:mm:ss |
| 9 | Time: Into Pre-Op | hh:mm:ss |
| 10 | Time: Prep Complete | hh:mm:ss |
| 11 | Time: Into Procedure Room | hh:mm:ss |
| 12 | Time: Anesthesia Induction | hh:mm:ss |
| 13 | Time: Procedure Start | hh:mm:ss |
| 14 | Time: Procedure Stop | hh:mm:ss |
| 15 | Time: Anesthesia Stop | hh:mm:ss |
| 16 | Time: Out of Procedure Room | hh:mm:ss |
| 17 | Time: Into PACU | hh:mm:ss |
| 18 | Time: Ready for Discharge | hh:mm:ss |
| 19 | Time: Out of Unit | hh:mm:ss |

We also collect the schedule that has the time of the case as planned in the morning and procedure room number, as seen in Table 2.

**Table 2:** information required from the schedule

| # | Data Element from Schedule | Format |
| --- | --- | --- |
| 1 | Case ID (or element to crosscheck with EHR) | Text |
| 2 | Procedure Scheduled Date & Time | mm/dd/yyyy hh:mm |
| 3 | Procedure Room Number | Text |
| 4 | Type of Procedure | Text |
| 5 | Preferred Provider (or ID) | Text (*anonymized*) |

To calibrate the digital twin, we run the simulation of a clinical day, starting from the schedule of that day, and computing the sequences of events from the registration of the first patient to the last patient leaving the GI lab. In each run, we use the projection operators of Figure 4 to compute statistical distribution of interesting metrics for the GI lab’s management – such as overall time spent by the patient in the GI lab, turnover time, time of first case, etc.& Each simulation alone are meaningless from a statistical point of view because the underlying discrete dynamical system is relatively small with a number of patients of the order of one hundred per day or less. We need to run each simulation of a clinical day enough times to get an overall distribution of these quantities close to convergence. The EHR data for a single clinical day suffers from the same limitation: the data set of a single clinical day is too small to produce a statistically meaningful estimate on turnover time, late start, overtime, etc. Consequently, the calibration of the digital twin is based on the fitting of these distributions on several days’ worth of data. In principle, the more clinical days the better, but the clinical conditions and infrastructure vary with time as well. We run the calibration of the digital twin on a time period of one month, at most, in order to limit the effect of staff turnaround. The optimization process’s search for the minimum of an objective function that is a weighted combination of the prediction accuracy on:

i. Distribution of the overall time spent by the patient in GI lab – see Figure 5 and 6.

**Figure 5:**
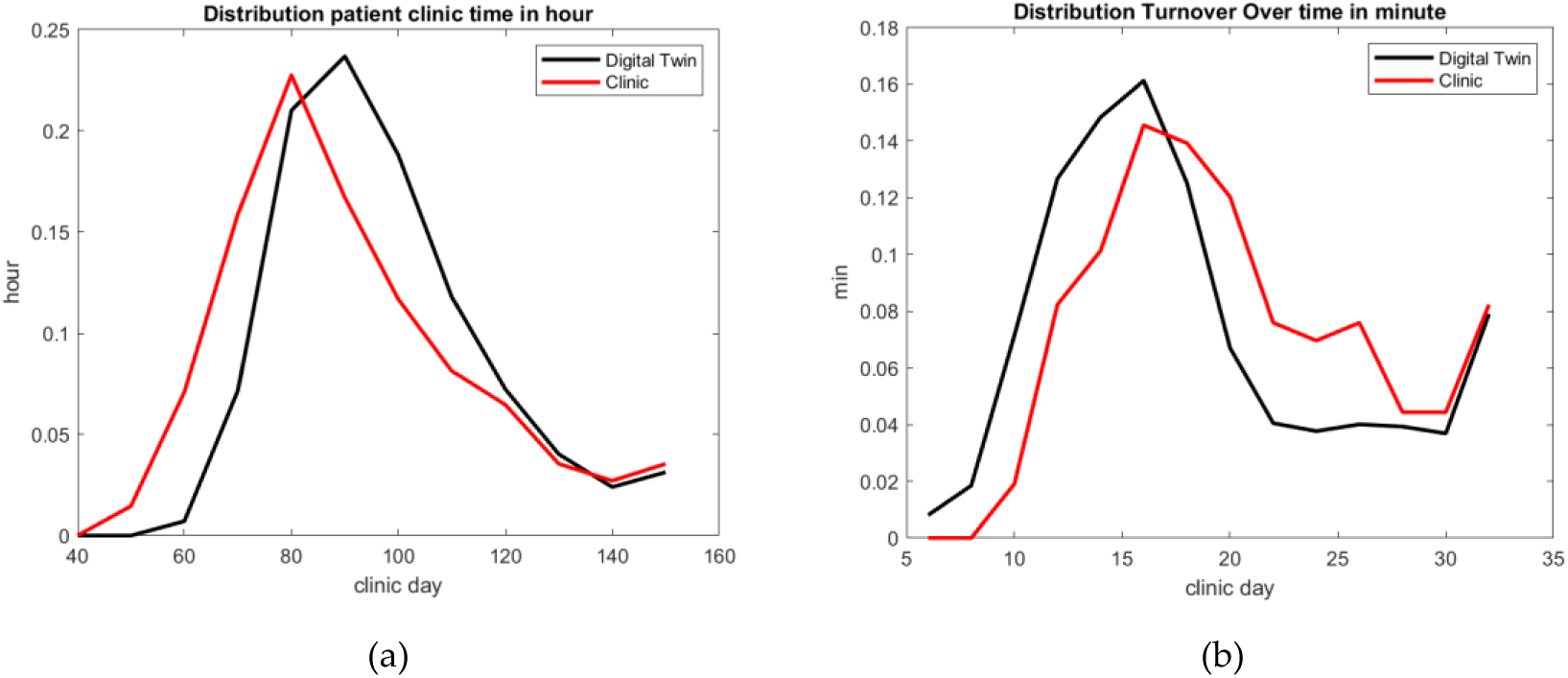
GI Lab 2: comparison of the best fitted digital twin model of patients’ total time in the GI lab (a) and turnover overtime (b).

**Figure 6:**
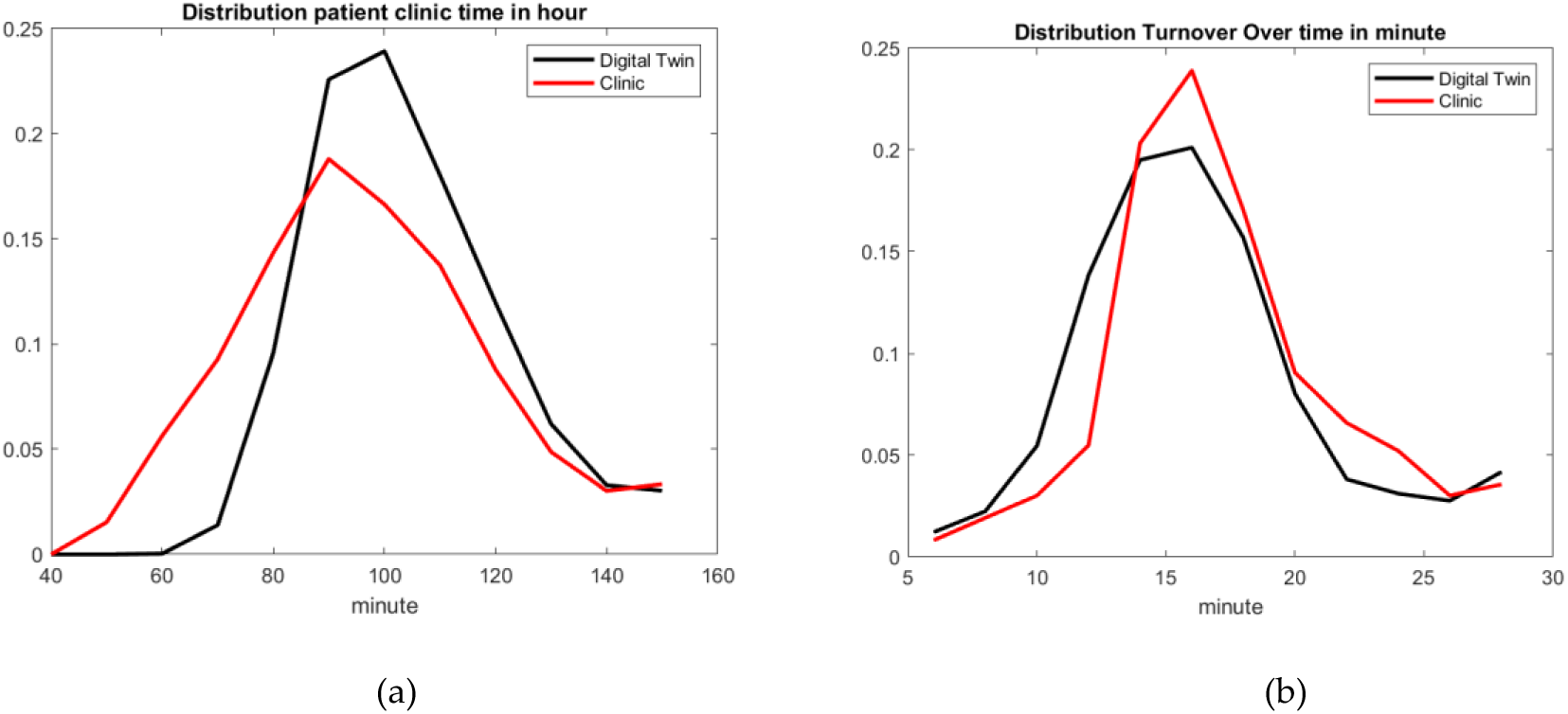
GI Lab 6: best fitted digital twin solution of patients’ total time in the GI lab (a) and turnover overtime (b).
ii. Distribution of turnover time – see Figure 5 and 6.
iii. Distribution of first case of the day start time in each procedural room.
iv. Distribution of last patient leaving the GI lab – see Figure 7 and 8.

**Figure 7:**
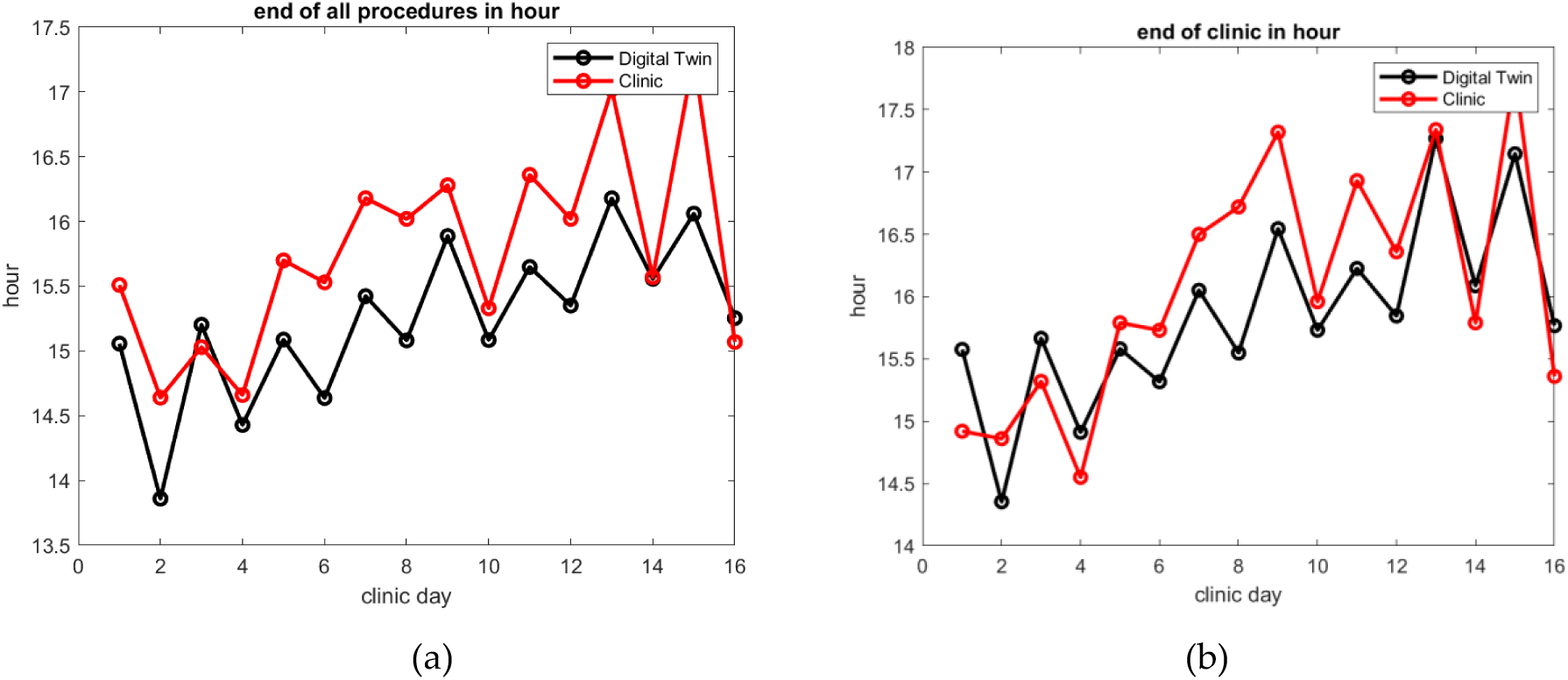
In GI Lab 2, comparison of best fitted digital twin prediction of the end of the last procedure (a) and the end of the clinical day (b). The average difference is 34 minutes for the end of the last procedure and 29 minutes for the end of the clinical day.

**Figure 8:**
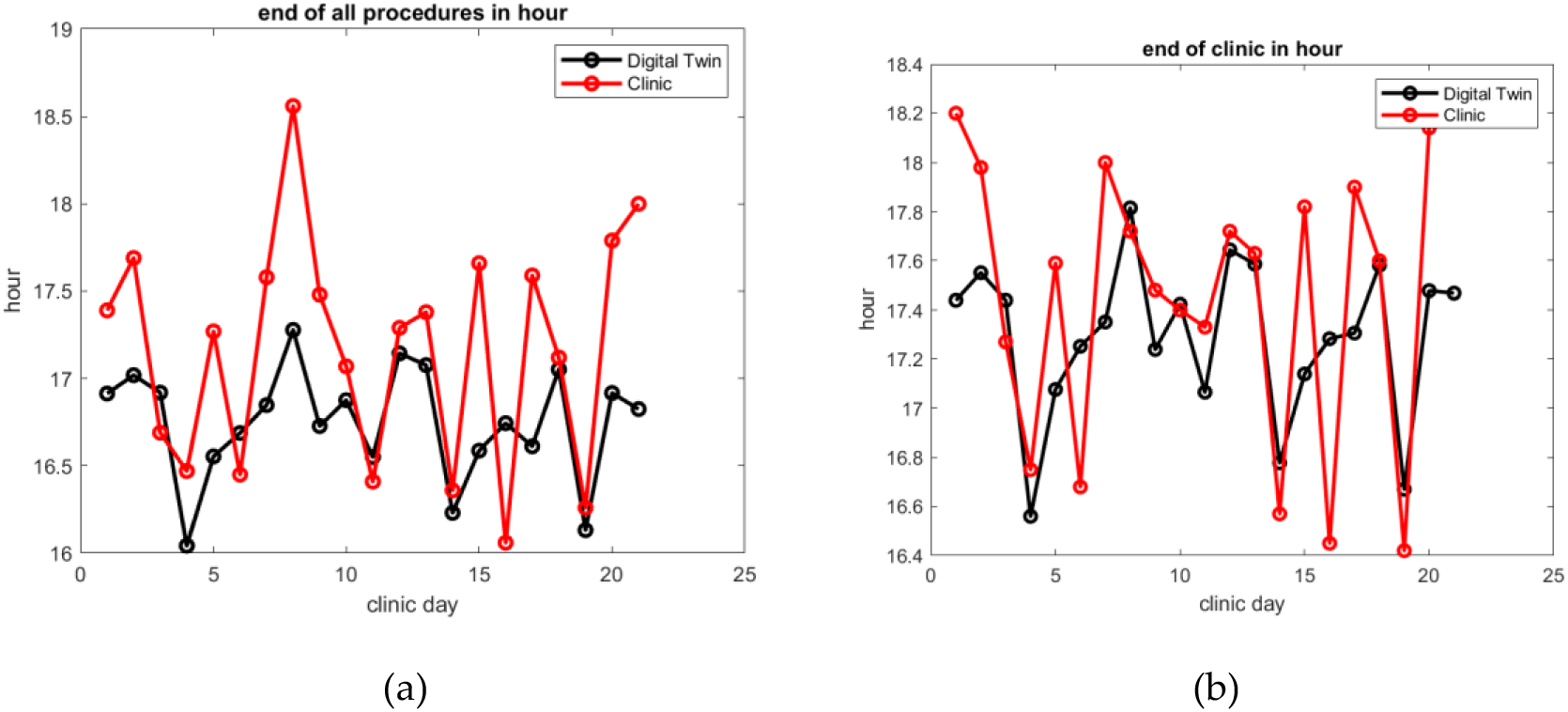
In GI Lab 6, comparison of best fitted digital twin prediction of the end of the last procedure (a) and the end of the clinical day (b). The average difference is 40 minutes only 80% of the time.

The objective function is constructed as follows: we compute the L2 norm of the difference between each distribution predicted by the digital twin and computed from the EHR timestamp data set. We normalized each of these error values by the L2 norm of the reference value. This produces a vector of four values per clinical day, one corresponding to each above criterion (i) to (iv). We average those vectors’ values over a time interval [0, T] that corresponds to every clinical day of the month. We take a weighted combination of all four entries of that average vector to get the function value to be minimized in the optimization process. This weighted combination can be decided as a function of what the management team likes to prioritize: an oversimplification would relate directly to staff satisfaction with (i) or productivity with (ii) and (iv), or staff satisfaction with (iii) and (iv).

The Formula (1) below summarizes the objective function; denoting D1 to D4 as each distribution listed above:

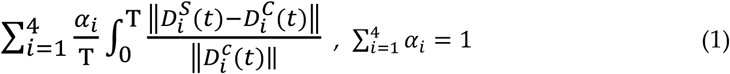

To solve the minimization process that leads to the optimum parameter set up of the digital twin, we use the stochastic optimization technique. We do not have access to the gradient of the objective function, and the digital twin is a stochastic process as well as a reality of workflow in the GI lab. The first technique that comes to mind is a genetic algorithm that can come in different mathematical formulations. It might also be desirable to track the objective function in the many forms that can take the weighted combination of Formula (1), to let the GI lab decide what could be a priority: Finishing the day early? Starting on time? Taking care of as many patients as possible?

We have run many scenarios and increased the robustness of our findings by keeping track of the objective function’s value using multi-objective results in every single evaluation of every stochastic optimization run. In the meantime, we found that a swarm algorithm can accelerate the convergence once the genetic algorithm has filtered out most of the parameter search space as non-competitive solutions [23].

At the end of this step in our method, a digital twin is constructed that is realistic enough to mimic any clinical day’s output of a specific GI lab with the same infrastructure conditions, in particular staff composition. This digital twin is specific to the GI lab for which we used the EHR data set. We can now use the digital twin to run many virtual experiments in an attempt to answer “what-if” questions for the GI lab.

In other words, getting answers on why delays accumulate under specific circumstances [17] or what would be the benefit of a new management decision, for example, can be put to the test in a computer algorithm. We will describe that process in the next section as an Artificial Intelligent (AI) method.

### 2.3 Heuristic Computer Reasoning supported by the Digital Twin

We are proposing now a HEuristic Reasoning ALgorithmic Development (HERALD) system that can support and inform the manager of a medical facility and greatly improve their efficiency and the efficacy of implemented systems see - Figure 5.

The starting point to reproduce this “thought experiment” [24] in silico is to:

- Generate a certain subset within a domain of hypothesis or questions and
- Simulate and trial said subset by running a “digital twin simulation” of the clinical workflow that integrates all the specificities of the GI lab.

To start with an example, a large amount of work has been done in scheduling based on optimizing the number of procedures on patients that can fit in a preassigned time grid [25,26,27]. One may do some assumptions on how long a procedure with a specific provider takes, based on previous records. The optimization process considers the infrastructure at a coarse level such as number of rooms, number of beds, and is very mechanical in nature. From the mathematical point of view, the optimization algorithm must work with a landscape that has multiple local optimums, described by a very noisy set of parameters: the success of the prediction of how long it takes to complete a medical procedure is very limited because it depends on many unknown factors [28].

What we propose is to run the schedule with the digital twin and observe what happens. If an optimization process of the schedule has any chance of success it should at least take all the system components into account, including a staff behavior description that has statistical meaning and multiple nonlinear interactions between the peri-operative area and the procedure room suite. Coordination delays between these different working units are captured by fitting the digital twin to the reality of the GI lab by design.

Now one can ask: will the clinical day end on time, or do I need to reschedule a procedure in a different room? If the latter, what would be the best rescheduling for that patient? Answering such complex questions is usually done by a manager who “knows the place” from experience and uses some intuition. With our methodology, we propose to improve the probability of the decision being the correct/best decision. To that effect, the digital twin simulation is running several simulations of the clinical day to predict the outcomes. Statistics on overtime or best room relocation is computed from that data set of outcomes and is delivered to answer the question of the GI lab’s manager. Trying many scenarios is a very heuristic way to answer questions, instead of a thought experiment that a manager does with intuition. It is an explicit algorithmic process that the computer can do systematically and quickly while considering all the known information entered in the digital twin. The decision might be only as good as the digital twin’s predictive capability is, but we enter here a rigorous process of validation that is well known in modeling. We will show in the results section specific examples of this in practice with true clinical data.

The architecture of the HERALD system is given in Figure 5. The best way to understand this representation is to show how this architecture can reproduce a digital version of “thought experiments” that are intended to structure “the process of intellectual deliberation within a specifiable problem domain” [14].

For convenience, we will take the classifications of Yeates [14] and illustrate each type of heuristic reasoning algorithm with an example of application on the surgical floor:

1. Prefactual: “What will be the outcome if event X occurs?”
  - What if the janitorial team is aware of procedure room state in real time?
  - What if the GI lab hires an additional anesthesiologist?
  - What if Provider X is late by 30 minutes?
2. Counter Factual: “What might have happened if X had happened instead of Y?”
  - What would have been the end of the clinical day in procedure room 6 if Provider X had been on time?
  - It is usually understood that nurses are specialized in Pre-Op or PACU: what if half of the nurses can be knowledgeable about various situations and work in multiple areas?
  - It is usually understood that a specific provider operates on their patient: what if a provider can treat any of the patients who have been scheduled?
3. Semi-Factual: “Even though X occurs instead of Y, would Z still occur?” We assume that the target Z is a component of the objective function:
  - Even though Nurse Y could not provide a necessary component to a surgical operation in time, does Provider X still finish in time?
  - Does productivity change with single-use scopes instead of reusable scopes?
  - Does the end of clinical day depend on the speed and efficiency of the procedure room turnover?
4. Predictive: “Can we provide forecasting from Stage Z?”
  - Can we predict the growth of revenue if we hire two more nurses?
  - Can we predict the patient satisfaction rating change if we overbook by 10%?
  - Can we predict end of the clinical day with high confidence at time X of the day?
  - Can we predict at 07:00. which procedure room will end the clinical day late?
5. Hind Casting: “Can we provide forecasting from stage Z with new event X?”
  - Can we assess how the next patients of procedure room 5 will be delayed since procedure X may take Y more minutes?
  - Patient Y has canceled, how much time of the day may Provider Z have free?
  - Duodenoscope Y did not pass the cleaning test after the washing cycle, do we anticipate a shortage of scopes for the ERCPs coming? What effect does this have on time and efficiency?
6. Retrodiction: “past observations, events and data are used as evidence to infer the process(es) that produced them”:
  - Procedure room 6 ended up finishing the day much later than expected, can we explain what were the main factors influencing the delay or delays?
  - Patient satisfaction has been declining this month, can we find out why this is different from last month?
  - There has been no more backflow around 11:00. in Pre-Op, what has led to this positive change?
7. Backcasting: “moving backwards in time, step-by-step, in as many stages as are considered necessary, from the future to the present to reveal the mechanism through which that particular specified future could be attained from the present”:
  - How early could we have predicted that procedure room 5 would end the day late by 2 hours?
  - How early could we have predicted that Provider X’s performance was impacting GI lab results?
  - How early could we have anticipated to release the staff early, or altogether, from their duties for the day?

We have shown in Appendix 1, that we can provide a mathematical formulation for each of these types of questions. This mathematical formulation corresponds to the architecture described in Figure 5 and is amenable to a solution with standard numerical analysis algorithms well known in data mining and optimization. The quality of the answer depends on the quality of input data that the digital twin receives. Questions in classifications 1 to 5 use multiple runs of the digital twin forward in time either from a given state variable or a modified state variable or a set of variables in a fixed neighborhood of variation of a given state variable.

Questions in classifications 6 and 7 requires solving the inverse problem on the state variable prediction using an optimization loop.

Since the digital twin is stochastic, we provide a probability estimate of the outcome as follows: if the answer is a numerical value, we should produce the average and standard deviation of the set of outputs corresponding to as many runs as needed to get within the confidence interval. If the answer is a discrete entity like “yes”, “no”, or “do not know”, we count the occurrence of each different answer from a large enough set of runs to get acceptable convergence.

A direct application of the process of digital thought experiments is to take action with predictive issue avoidance. HERALD first needs to automatically generate certain questions whereby the basic philosophy is to continuously investigate what may go wrong (and/or correctly) in order to bring remedies before an event even happens. For now, we may rely on a database of frequently asked question from the GI lab’s manager, but a further AI task is to generate the most relevant questions with an algorithm that mimics curiosity and learns from mistakes [29]. The system will then be able to ask the right question at the right time and give objective information to the manager on what will happen during the clinical day.

Next, we will describe the results we have obtained with our method to understand workflow management improvement in an outpatient center [30].

## 3. Results

This section may be divided by subheadings. It should provide a concise and precise description of the experimental results, their interpretation, as well as the experimental conclusions that can be drawn.

### 3.1. Data Set of our clinical studies and Evaluation of unintended manual error input

We have collected data from six outpatient GI labs named 1 to 6, over several months as summarized in Table 3.

**Table 3:** Distribution of the number of patients present in the database for each GI lab analyzed. Each center has different number of procedure rooms and different distributions between the two most common GI procedures or a combination of the two (double).

| GI Lab Name | 1 | 2 | 3 | 4 | 5 | 6 |
| --- | --- | --- | --- | --- | --- | --- |
| Number of Procedure Rooms | 7 | 3 | 3 | 2 | 2 | 7 |
| Colonoscopy | 6916 | 7339 | 5657 | 3261 | 1335 | 4848 |
| EGD | 1827 | 2299 | 2593 | 1792 | 794 | 1731 |
| Double | 1520 | 1945 | 891 | 161 | 404 | 1489 |
| Time Period | Aug-Dec-2018 & Aug-Oct -2019 | 2019 | 2019 | 2019 | 2019 | Aug-Dec-2019 |

The volume of procedures per procedure room per year is shown in Figure 1. It shows as stated in the background section (iii) that every GI lab has very different performances.

Since we want to use this data set to calibrate the digital twin, we need to assess the quality of the data sets. It is important to notice that the rate of error in both data sets is not negligeable, see Figure 2.

To start, we observe missing entries in the EHR’s raw timestamps per procedure; second the timestamps can be out of order, which is impossible since the workflow must go forward in time from registration to discharge, except for anesthesia that may start in Pre-Op and stop in PACU, not respecting the order of Table 1.

This is not very surprising because all this information is entered manually by medical staff. Typically, nurses are focusing on their patients and, depending on conditions, may either forget to enter the timestamp information or enter it after the occurrence, a manipulation that EHR systems do not handle very well. We found as illustrated in Figure 2 that 1 out of 5 procedure’s records is usually incorrect. Figure 3 gives some insight on the fact that manual entries are impacted by human behavior and fatigue. The error rates have the tendency to increase as the morning goes, decrease after the lunch break, to increase again as the end of the clinical day gets closer.

There can be also a significant number of incorrect entries on procedure room number allocations in the schedule, about 10% of these records are incorrect in GI Lab 6 and detected when there is overlapping times of procedure on different procedures in the same procedure room. Overall, as stated in the background section (ii), EHR entries have significant inaccuracy that make workflow optimization more difficult to achieve.

To overtake some of these limitations, we have implemented in two of the outpatient centers, with the largest volumes, a cyber-physical infrastructure that collects the workflow timestamps automatically. Most importantly, we acquire very accurate and reliable information on procedure start times and end times, by grabbing the endoscope video output in real-time to detect when the scope is active or not inside the patient or outside. We refer to our previous work published in [30] for more details. We observe that EHR timestamps of procedure start and end may shift task events by a few minutes randomly in 10% to 50% of cases.

For comparison purposes, we use a second data set where the input entries on procedure timestamps are obtained by analyzing the endoscopic feed in real-time with our computer vision algorithm. This implementation was done only in GI Labs 2 and 6. This second data set has 1944 patient records in GI Lab 2 and 10412 patient records in GI Lab 6. We found that the techniques in our cyber-physical infrastructure implementation are more reliable than the manually entered EHR timestamps and capable of correcting many incorrect EHR entries [30]. This analysis confirmed the procedure room number errors as well. By looking at the start and end timestamps of procedures in the EHR and the ones from our computer vision solution, we can accurately reconstruct what happened in the procedure rooms and what the exact order of procedures was in each procedure room, see Figure 11. This solution doubles in value as it also gives us more information on the scope usage than what can usually be found in the EHR system. In this second data set, however, manual EHR entries of perioperative timestamps are still used and suffer from similar inaccuracy than in the first data set of Table 3.

Finally, as noticed in the background section (ii), we have seen seasonal effects on the number of patients showing up for their medical procedure, and change in staff composition over the year, such that the clinical workflow performance may vary greatly from one month to the next. Next, we will present the calibration of the digital twin of GI Labs 2 and 6, where we have the most experience to interpret the results.

### 3.2. Result on the calibration od the digital twin

We found the optimization process, described in our methodology section, is robust provided that we run at least 20 times each day simulation. The overall calibration process takes approximatively 12 hours on a standard PC desktop equipped with an Intel processor (11th Generation Intel i7, 4 cores/8 threads with 16GB of RAM). We can accelerate the code via parallel computing: by running in parallel the set of parameters for each generation of the combined genetic algorithm and/or swarm algorithm[32]. The speed up of such code is known to be close to optimal, i.e., the acceleration factor is close to the number of processors used.

As shown in Figures 5 and 6, for GI Labs 2 and 6 respectively, the digital twin reproduces turnover time with good accuracy. Turnover is defined here as the time between scope out of one patient to scope in of the next patient. Since we used our computer vision solution to detect these events, the distribution of procedure turnover time is accurate. Turnover is one of the most nonlinear output variables [17], since it depends on so many events such as patient is ready, provider meeting next patient in Pre-Op, provider meeting prior patient in PACU for discharge, scope ready, Certified Registered Nurse Anesthetist (CRNA) ready, etc.

The digital twin reproduces the overall time spent by a patient in the GI lab with satisfactory accuracy, see Figures 5 and 6. It should be noticed however that the EHR record gives unrealistic short time presence for some patients. In reality, it is impossible for a patient to get through the whole GI care process in less than 60 minutes. These EHR incorrect manual entries impact the distribution on the left side of the bell-like curve.

Late starts depend on many unknowns related to the clinical team, equipment preparedness, and other factors in external activity. We verified that the statistical model used in the digital twin to provide first procedure starts has the same level of relatively uniform randomness observed in the clinical data. We also found from simulation that this variable does not have lead impact on the validation.

We found that matching the distribution of overall time spent by a patient in the GI lab (i) and matching the distribution of turnover time (ii) provides enough information to calibrate the digital twin. In other words, adding (iv) as an objective criterion does not make any improvement in the fitting. Therefore, (iv) can be used separately for validation.

### 3.3 Is the digital twin useful?

We have tested our digital twin against some of the questions heuristic computer reasoning may address. The first question is about predictive ability: can we provide forecasting of overtime? Can we predict at 07:00 which procedure room will end its clinical day late?

Figures 7 and 8 show the capability of the digital twin to predict the end of the clinical day for all patients, as well as the end of the clinical days for all providers. The prediction is significant if the error is less than 30 minutes, which is the standard duration of a block in scheduling. This prediction holds true only if the error rate on room allocation in the schedule is moderate, i.e., about 5% error. The result in GI Lab 6 is not very accurate, but if we restrict ourselves to days with less than 5 % error on room allocation in the EHR record, i.e., 8 clinical days out of 21, we found that the prediction on end of clinical days has an average accuracy of 23 minutes. We expect that this prediction can be adjusted as the day advances to help staff anticipate when they can leave the GI lab.

We also indirectly tested our digital twin against a counterfactual question: What would have been the end of clinical day if cancelation rate were to drop to zero?

We run a simulation of the digital twin testing various cancelation rates virtually and observed the impact on the prediction of the end of the clinical day: we observed that a range of 2% to 8% cancelation rate has virtually no effect on the end of clinical day. To compare that finding to a true observation in the GI lab, we examine our data set with two different months that have similar workloads and different cancelation rates. We noticed that while the cancelation rate varies from 2% to 8% over these two months, the times for end of clinical day stayed about the same.

The true observation of the GI lab is therefore similar to the “virtual observation” provided by the digital twin in the same range of cancelation rates. We interpret the result as follows: when cancelation occurs, one needs to reschedule the next patient(s) in real-time; but to do so efficiently one needs first a real-time awareness on the workflow in the GI lab, second a very quick management decision and coordination with the various staff involved in the workflow. We found that those two conditions are difficult to achieve simultaneously in practice, with the current process of GI Labs 2 and 6 that both rely exclusively on manager experience and intuition.

We run a third test of a similar nature: what is the impact on the GI lab if patients spend 6 minutes less in the pre-op area? The rational for this question comes from our experience during the installation of our cyber-physical infrastructure in GI Lab 6. We influenced improvement in their workflow efficiency by installing a new graphical user interface in the pre-op area to increase awareness of the nurses and improve communication with busy providers. Over a one-month period, we observed a net decrease of the time spent by the patient in the pre-op area by 6 minutes. Eventually the GI lab’s throughput increased by 11% without increasing the total amount of hours worked by the staff [30]. The comparison between the two data sets before and after the 6-minute gain allowed us to test the heuristic computer reasoning algorithm addressing the following question: could we have predicted this improvement on throughput? It turns out that the digital twin predicted that the correct number of working hours freed by the 6-minute gain in pre-op allows the GI lab to take care of 10% more patients. Such a benefit seems very significant compared to what a 6-minute average saving per patient in preparation time seems to be. In practice, GI Lab 6 was very efficient to start with and initially managed to take care of 14 to 15 patients per procedure room per clinical day. It is not surprising then that a small change in pre-op duration has a great impact on performance.

#### Retrodiction

can we detect what are the main factors influencing overtime? This question is easier to answer in principle and can be addressed at first by a linear sensitivity analysis. Saving time in the pre-op phase seems to be the main factor, while saving time in the PACU phase comes second. The effect of accelerating turnover seems less important. We have unfortunately not been able to test these last two results against clinical data yet. It would have requested a change in staff performance that we have not been able to observe so far in our clinical study, and the result might be different from month-to-month as the GI lab improves its workflow components.

According to our initial findings, global prediction involving metrics on a large time interval, and therefore affecting many events and patients, seems to work well when the digital twin is calibrated on the EHR data for a monthly period. It is less clear to pinpoint the impact of specific individual behaviors during a clinical day. Examples such as the following questions would require accurate tracking of events during the clinical day and better manual entries:

#### Prefactual

What if provider X is late by 30 minutes?

#### Semi-factual

If colonoscopy duration of provider X has no more overtime, would that make a difference on overtime?

#### Hindcasting

Can we assess how long the next patients of procedure room T will be delayed since procedure X may take Y more minutes?

Any error in EHR timestamp entries or incorrect room allocation in the schedule specific to that individual event, would ruin the accuracy on the initial condition that the digital twin needs to run. Therefore, we are continuing to develop a cyber-physical solution that captures the perioperative workflow independently of the EHR system [30] to accompany our cyber-physical infrastructure presented in this paper. Its impact can be better understood by examining how a change of schedule is handled during a typical clinical day, as shown in next section.

Let us now discuss the potential use of the digital twin to improve scheduling as the clinical day progresses. We will present this result as a conjecture to open some perspective: our goal in a future publication is to get the acceptance of a GI lab to use the output of our new real-time rescheduling solution and check its efficiency.

### 3.4 Conjecture

It is well known that schedules rarely go as initially planned in a surgical suite. This is also true for GI outpatient centers. Figure 9, for example, reports on the number of scheduled changes per clinical day over a period of one month in GI Lab 2. In practice, a change in schedule is defined as a block being removed, a block being added, or a shift of one block by 30 minutes or more.

**Figure 9:**
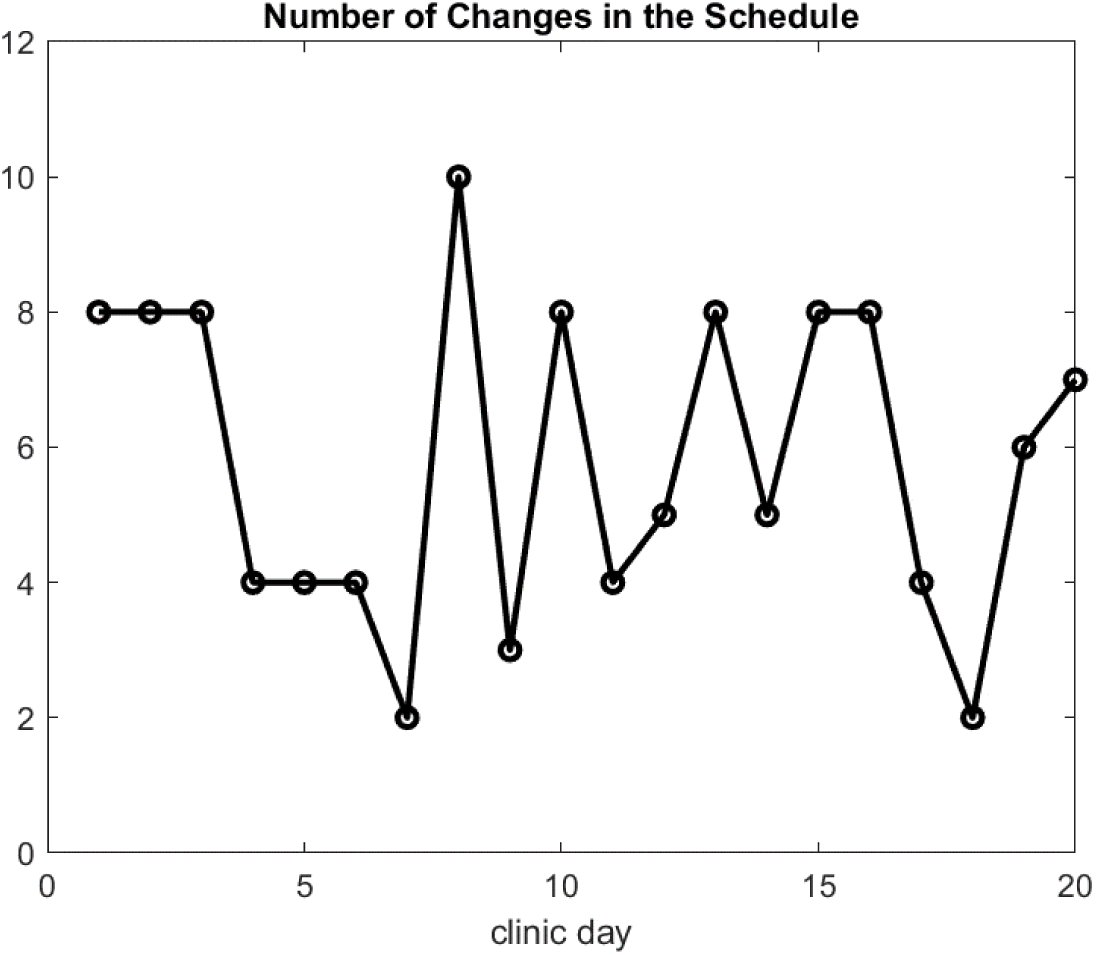
Number of changes in the schedule per clinical day due either to cancelations, add-ons, or very late patient arrivals.

Scheduling in GI outpatient centers is a simple matter: the common practice is to allocate a block of 30 minutes to every patient no matter if it is an EGD, a colonoscopy, or both (double). The procedure is supposed to start at the hour or half past the hour. Figure 10 gives two examples of block scheduling in four procedural rooms, with one not being used. The scheduling on the left is the one implemented by the GI lab that day. Blocks are color coded according to the type of procedure: red for colonoscopy, blue for EGD, green for double. The two letters in each block identify the provider. Scheduling is often done manually or by software with additional manual entries. The first block of room 1 in the left, initial schedule of Figure 10 was unfortunately allocated to two different providers. This type of error may come from a late manual entry that did not update the scheduler properly. The visualization of the schedule is then incomplete with procedures being hidden behind others due to an error in room number or scheduled time of the procedure. This error usually delays the end time of the clinical day, forcing staff to work extended hours and possibly overtime.

**Figure 10:**
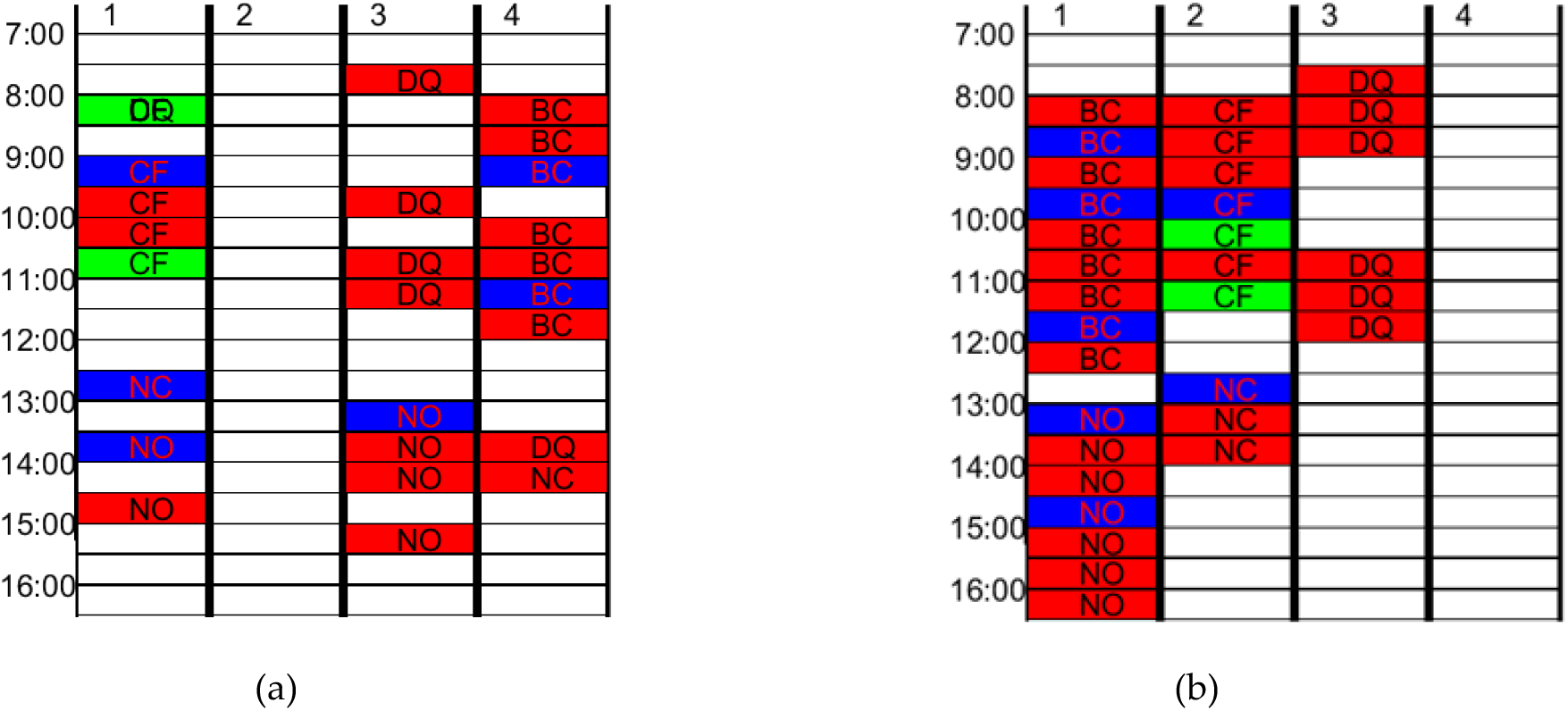
Optimizing rescheduling in real-time while making sure that patient will be done with their preparation in due time: (a) clinical day as it happens; (b) optimum rescheduling.

Fixed block scheduling is common practice: all 6 outpatient centers that we have documented data from in Table 1 use the same scheduling assumption. It does not count for the fact that EGD procedures finish sooner than a colonoscopy and a double procedure would. It is true that this difference does not really matter, provided that there is a good mix of these three procedure types over the clinical day in a procedural room (one shorter procedure may compensate for another that takes longer). Still, the schedule hardly represents the reality.

Figure 12 gives a representative example of what happens in GI Lab 2 when decisions with rescheduling are made based on the intuition of the manager or any staff who may have a say. As we described earlier, our cyber-physical sensor (termed as our FES sensor) installed on the endoscopic video system allows us to have automatic timestamps on usage, state of scope (i.e., scope active, scope out but plugged, etc.), identification of scope reference number, and type of scope. There are instances where two different scopes are used during the same procedure. From this information consolidated with the EHR system, we create an accurate timeline of procedure room states – see Figure 11. The classic order is the patient enters the room (black bar with a circle on the graph) then the procedure starts (red block for colonoscopy/lower, blue for EGD/upper) and last the patient leaves the room (black line with a triangle) - double procedures can be recognized as the quick succession of an active upper scope followed by an active lower scope. We can also compare this timeline corresponding to the true procedural room utilization with the timeline corresponding to the block scheduling of that same procedural room as initially scheduled before the clinical day started. By overlaying these two timelines, following a logical order and matching start and end of procedures, we can visualize when there is a match or difference between block allocations, as exemplified in Figure 12. Differences can be of different nature: (i) missing procedure in the EHR, (ii) different order of procedures and last, but not least, (iii) wrong room number. To point out these differences with accuracy, we have shown on the right of Figure 12, the initial schedule at the beginning of the day with 30-minute block allocation per patient. According to the initial schedule, in Room 1 provider ‘NC’ is supposed to have 10 colonoscopies. The reality was different, first another provider called ‘MF’ had one procedure in this room that day. This colonoscopy was scheduled but scheduled for the same time as NC’s 3rd colonoscopy at 09:00. NC performed 12 procedures that day instead of the 10 according to the initial schedule. Indeed, two other procedures, an EGD and a double, were scheduled at the same times of two colonoscopies (one at 13:30 and one at 14:00). Because of this, the staff in this room finished at 15:30 rather than at 14:30. A similar event happened in Room 2, with provider DH having one EGD and one colonoscopy scheduled at 10:00 making the colonoscopy named ‘DH8’ on the timeline start at 12:15 instead of 10:30; overall DH is finishing the day one hour late as well. One can wonder, for example, why MF did one procedure in Room 1 while there were free blocks available in Room 4 where MF’s other procedures took place. Now, we will present a software solution to achieve rescheduling in real-time that can be tested with the digital twin prior to making any decision in order to avoid missing opportunities in procedural room allocations.

**Figure 11:**
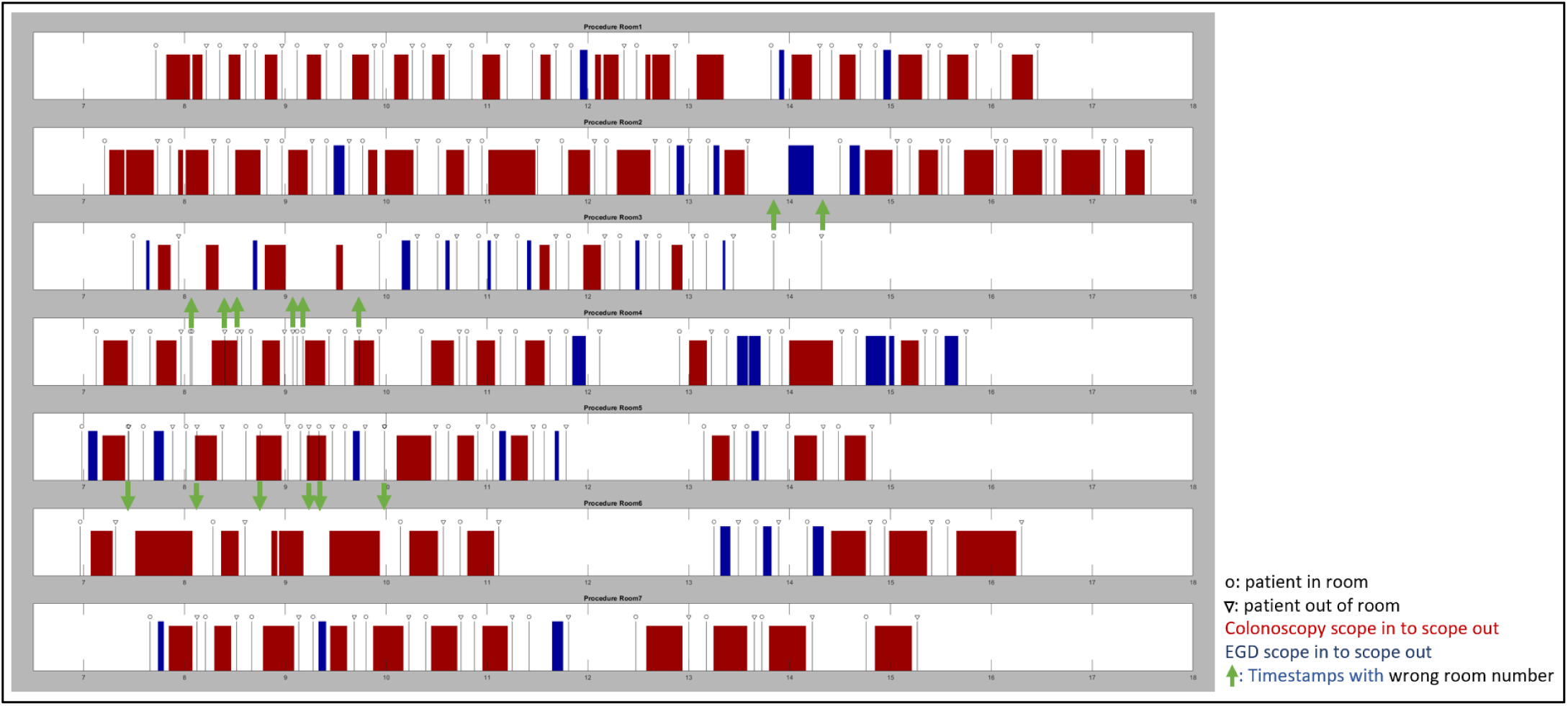
An example during one day, in one clinic with 7 rooms, from 07:00 to 18:00 of output from the EHR data (black line with circle and triangle showing when the patient enters and leaves the room respectively) and from our computer vision systems (red and blue blocks showing when the scope (lower in red, upper in blue) is active; i.e., procedure is ongoing). The green arrows show when there is an error of room number and where the EHR timestamps should actually be.

**Figure 12:**
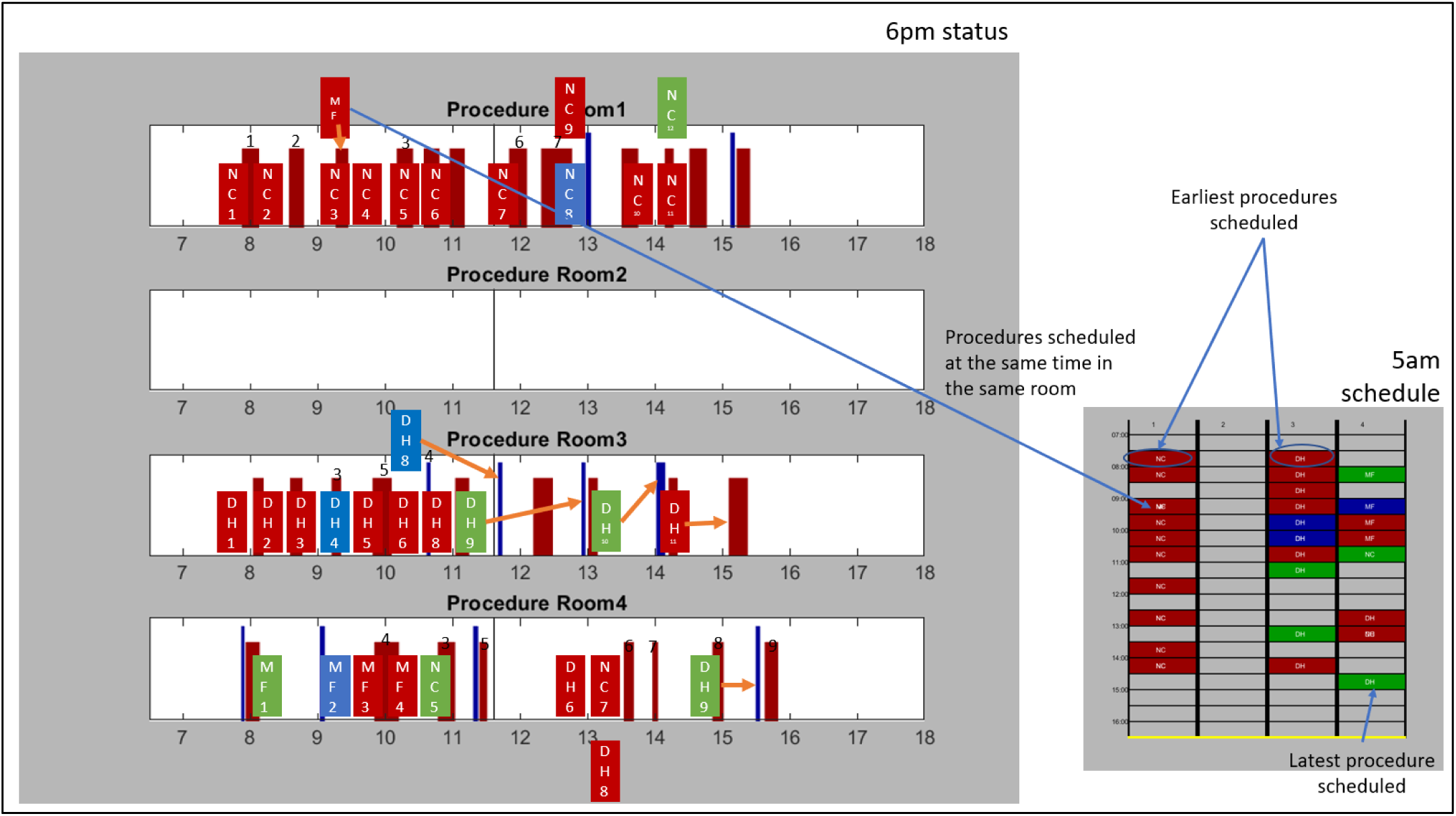
An example of missed opportunities during the rescheduling on a given clinical day and representation of the impact of double booking on the same block. The blocks with initials in them represent the scheduled time of procedure, following a 30-min block scheduling rule. The red and blue blocks behind them are the output of our computer vision system (like in Fig. 11) and shows the real time of start and stop of the endoscopic procedure. The orange arrows show when there is a large difference between the initial schedule and the reality.

As a first example, Figure 10 shows the schedule as it was executed on the left and the schedule that our real-time rescheduling software would suggest on the right. We will describe first the algorithm and second the benefit according to the digital twin simulation.

To improve the rescheduling in real-time, we first need to consider when the patient registers: there is no point to schedule a procedure and assign it a room unless one makes sure that the patient has plenty of time to go through the registration process and the pre-op phase before moving to the procedure room. We reorder patients in the schedule from the same provider to satisfy this constraint. Second, no matter how early a patient may show up, it is not generally accepted to ask the provider to start the day earlier. We conclusively impose the same start time of the clinical day in each procedure room as it is listed in the initial schedule. Third, we privilege the solution where a provider stays in the same procedure room to minimize loss in time due to moving from one place to another. It is relatively simple to use a standard bin-packing algorithm to build the schedule with only three procedure rooms [33]. As the number of procedure rooms increases, the algorithm may fail. One usually runs the search algorithm several times and produces several solutions that are local optimum of the optimization problem. In the example of Figure 10, one can conclude visually that the rescheduling improves the usage of each procedure room and should improve the rate of room occupancy. A rational way to choose the best solution would be to use the digital twin simulation output to look at multiple factors, such as turnover performance, overtime, end of day prediction, and patients’ and staff’s satisfaction.

To test this hypothesis, we have applied our real-time rescheduling virtually over a period of one month corresponding to the data set used to plot Figure 7. The result we found is summarized in Figure 13. According to this graph obtained with the digital twin, one can add about 5 procedures per day, with no change on scheduled start time of providers and neither assuming any increase on performance of the staff since the parameters that define the digital twin have been fixed. A quick estimate predicts that it would be possible to add 100 more patients per month without increasing the end time of clinical day neither the number of staff, which should result into an increase on gross revenue of about $100K per month. However, the validation of this rescheduling method needs to be tested out in true clinical conditions carefully and is the goal of another future publication.

**Figure 13:**
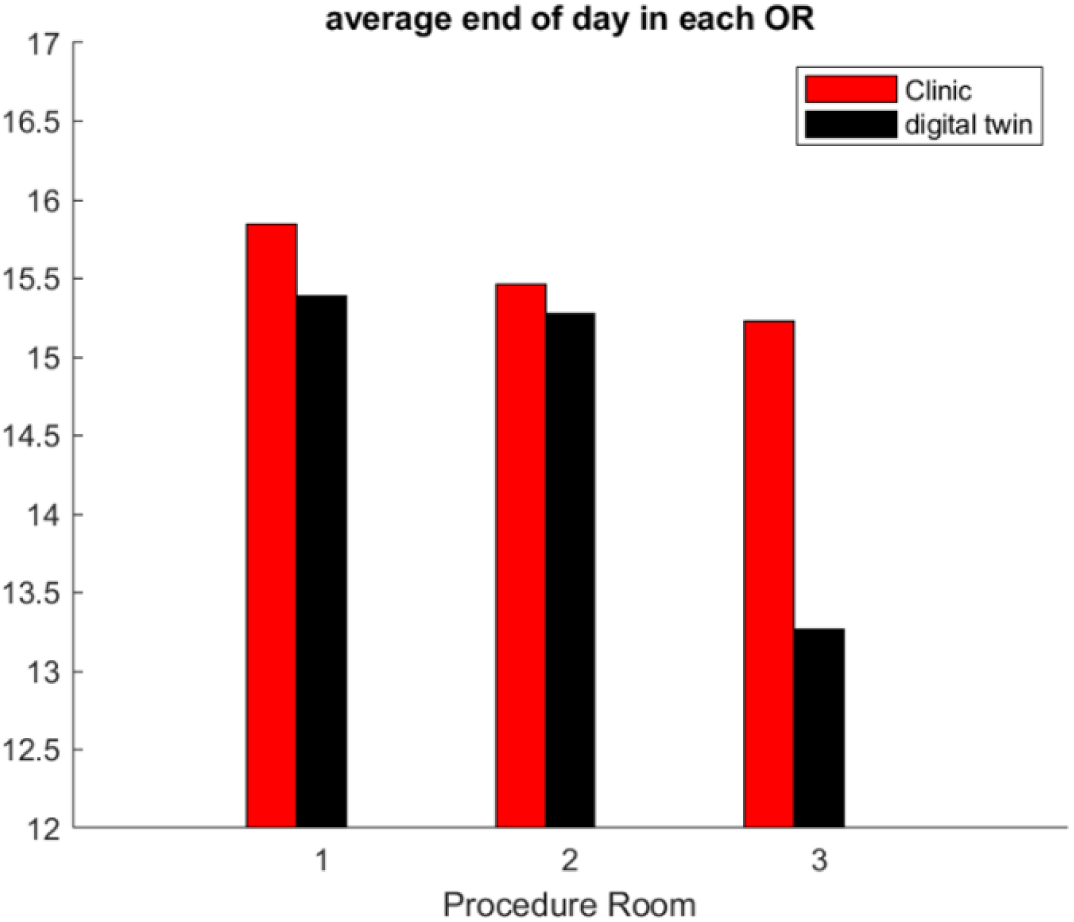
Impact of optimum rescheduling over a month based on end of clinical day in each of the three procedural rooms, according to the digital twin.

Next, we will discuss our results and conclude our study.

## 4. Discussion

We have concentrated our study on understanding how optimization of workflow in GI outpatient centers can be achieved. This is an important problem to solve: according to the Beckers Healthcare 2019 report [34], GI endoscopies make up for 68% of all endoscopies in the U.S., and GI procedures are estimated to grow at a 2.6 % compound annual rate, driven by the increased aging demographics and growing prevalence of GI-related conditions.

While the workflow in GI labs is much simpler than in multi-specialty hospitals, we found that workflow optimization is still an unsolved problem for the most part, as illustrated by the ASGE benchmark report [11]. Every GI lab was developed under different healthcare environments and standards. The GI outpatient center market is going through a strong phase of consolidation and merging [35] that gives hope for a more systematic way to establish a standard of optimum workflow and safety. However, it is difficult to generalize a solution that is intended to work for all GI labs. Besides, the data used to understand the problem come essentially from the EHR of the GI lab that can include a multitude of software platforms with poor interconnectivity: not only may the data be difficult to reconciliate but as shown in our study data may come with a high rate of error too.

To render the process of data collection unbiased and automatic, one may argue that a cyber-physical infrastructure like the one we developed in [30] might be a solution. However, we still need a solution to answer the most frequent questions of workflow optimization that are asked by managers and staff. Some, for example, were listed in our methodology section. Answering these questions in real-time to assist the management’s decisions might be difficult with today’s EHR data input quality. We have shown that the ability to predict the end of the clinical day with our digital twin model is feasible if the rate of error in the initial schedule is low.

Some more fundamental questions on strategy and resource allocation that rely on a larger data set, which looks at many days of past activities and prospects of new management solutions, might be easier to deal with. Unintended manual entry errors add random noise with unbiased distribution. For example, we found that the time shift on procedure start and end times due to manual entry approximations had no bias that would favor early or late timing.

## 5. Conclusions

Rather than using machine learning or deep learning methods that extract patterns in workflow timestamp data, we have chosen to use a digital twin of the GI lab that implements a priori knowledge on how workflow is bounded and should operate. This type of heuristic computer reasoning may deliver an explanation about why things turned bad as opposed to pure observation. For example, we were able to test the influence on preparation time of patients and show how it can be a major bottleneck on patient workflow in a GI lab that was otherwise fairly efficient. We were also able to verify that preparation time can be improved without impacting the PACU phase later down the workflow path. The same result would not hold true in a GI lab that has very poor coordination between staff in each phase as reflected in the study of the rescheduling in real-time by the digital twin simulation.

Another limit of standard AI algorithms in machine learning and deep learning applications might be in the size of the data and the error rate. A standard facility that treats about 1000 patients per month gives only 12K data points with a 10% to 20% error rate, at least. Unless one combines several centers’ data sets with the same exact infrastructure and number of shared staffs, the data set may remain far too small to support standard AI methods. On the other hand, the digital twin needs the adjustment of a few dozen of parameters at most, which seems easier to achieve with a stochastic optimization algorithm that naturally filters out the noise in the data set and gives useful results as seen in our two GI labs examples.

There are still strong limits to our methods. There might be numerous sources of singular events either due to the peculiar behavior of some individuals or some hidden rules related to staff scheduling and rotation that might be missed in the digital twin. Continually comparing the outcome of the digital twin with the realizations of the workflow in the GI lab on a regular basis may help to resolve these difficulties. It seems that on-site GI lab visits done in collaboration with a human factors team would be the ultimate way to improve and validate the system further. We will report on this activity in a later publication. Improving data input in our model both in quality and in finer time scales might also be important as the technology matures.

Finally, we would like to observe that the same framework might be implemented in other clinical workflow specialties such as ophthalmology in “laser eye surgery clinics”, or minimally invasive vascular intervention in catheterization laboratory. Each of these workflows share the same features as seen in GI: the workflow path is well defined from registration to discharge, medical staff all along the workflow path of the patient have well defined healthcare to deliver, procedures are relatively standard and short in time. From the mathematical point of view, the optimization workflow problem is “well posed”: our methodology applied to GI outpatient centers should be an effective tool to advance the field in these other clinic specialties. Our goal is to provide a tool that can inform data-driven decisions to improve both efficiency and safety in similar centers and improve our basic knowledge of what is the optimum path for the patient.

## 6. Patents

A provisional patent has been submitted “Architecture of a Heuristic Computer Reasoning System” Provisional 63171824.

## Supporting information

ethicDterminationLetter

## Data Availability

All data produced in the present study are available upon reasonable request to the authors except if asked otherwise by the clinics.

## Author Contributions

This project is highly interdisciplinary and required all co-authors’ contributions to establish the concept and reach the goal of the paper. All authors contributed to the writing of the paper. M.G. led the project, designed the overall mathematical framework, including the heuristic reasoning model, and Matlab code implementation. G.J. participated in the validation of the method and verified the experiments. S.F. participated in the gathering and analysis of the data from different EHR software. All authors have read and agreed to the published version of the manuscript.

## Funding

The pilot studies where the data come from and where the system was installed was funded by the company Boston Scientific Endoscopy. This funding was provided to the company ORintelligence, LLC with which the authors were affiliated, not to the authors personally.

## Institutional Review Board Statement

Not applicable

## Informed Consent Statement

the study did not involve humans.

## Conflicts of Interest

The funders had no role in the design of the study; in the collection, analyses, or interpretation of data; in the writing of the manuscript, or in the decision to publish the results.

## Appendix A

### Digital Twin and Heuristic Reasoning Abstraction

#### 1 Mathematical Framework

We introduce the following mathematical framework to implement the digital twin and algorithm of heuristic experimental reasoning:

- 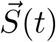 state of the system at time *t*.
- 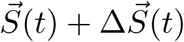 new event that updates state variable at time *t*.
- 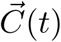 control variable at time *t*.
- 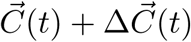 new change on control variable at time *t*.
- Observation that tracks the state variable evolution in time for a given control variable:

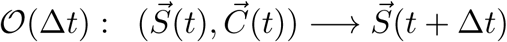
- Simulation algorithm that predicts the state variable evolution in time for a given control variable.

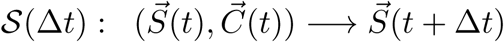
- Projection of the state into the objective value space is denoted

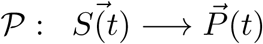 The projected value is a multidimentional vector (*P*_1_, …, *P*_*N*_).
- The objective function is the weighted norm:

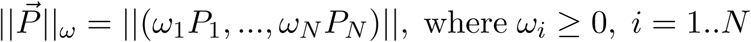
- Measure of the difference between realization and simulation on objective value:

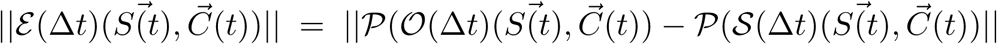
- continuity of error estimate with respect to state variable:

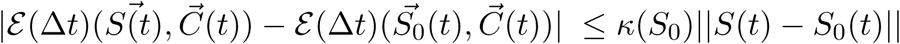

where *κ*(*S*_0_) denotes a real number that depends only on the state value *S*_0_.
- continuity of error estimate with respect to control variable

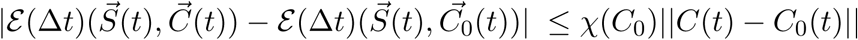

where *χ*(*C*_0_) denotes a real number that depends only on the control value *C*_0_.

Let us describe the state variable of the GI workflow:

##### State Variable

We keep track of the four following components of 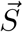 that are respectively procedure state, procedural room state, patient state and medical equipment state; Table 1 gives in each column an example of the meaning of each state value component corresponding to a GI lab. We define *S*_*j*_, *j ∈* (1..4) to be the row number that corresponds to the state of that component.

**Table 1.** procedure state, room state, patient state and endoscope state

| row # | Procedure | Room | Patient | Scope |
| --- | --- | --- | --- | --- |
| 1 | No Scope | Empty | not arrived | in storage |
| 2 | Scope Plug | Busy | at registration | Transport to Procedure Room |
| 3 | Scope In | Cleaning | in Prep | in procedure room |
| 4 | Scope Out | | in Procedural Room $x$ | Transport to Tech room |
| 5 |  |  | in Pacu | Hand Wash |
| 6 |  |  | Discharge | Washing Machine Cycle |
| 7 |  |  | Left the clinic | Drying |

We may fill up some additional row in this table with more detailed work flow steps on demand. We can also have additional vector components to capture other dimensions of the workflow. For example, we download relevant information on weather forecast and traffic condition to estimate the impact on inflow/outflow of patients in an outpatient center. We can keep track also of the mental state of staff by analyzing behavioral data such as response to text messaging, usage of touch screen, impact on task time line after text messaging, etc… We refer to the foundation work of Rosalind Picard on affective computing [8] for that purpose.

##### Control variable

Control variable are referring to the resources of the infrastructure allocated to the workflow: 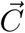 has the number of staff in each category, number of procedural room open, number of scope available for each category, number of stretchers, information sent etc…

The manager controls these variables within specific boundaries: for example, the manager may decide on the number of nurses allocated to prep and pacu based on the schedule load. To save on infrastructure cost, the manager can close one procedure room for half a day, if the number of procedures scheduled at that time allows it.

##### Projected Value

The outcome of the workflow is evaluated as a function of: turnover time between procedures, time spend by the patient at each stage of the workflow, economic output based on reimbursement and cost of infrastructure and staff, overtime, quality of service based on clinical outcome, etc… For each of these fields one may look at average value, standard deviation over a period of time. Each of these numbers can be weighted out to define a multi-objective function 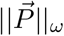, that we named the objective value.

##### Description of Workflow and Abstraction with a Model

The workflow is described by a directed graph *𝒢* illustrated in Figure 3.

Each node of the graph represents a step of the process or a task.

The task advances in time provided all conditions to start that process step are met. Those conditions are linked to patient state and depend on specific resources such as staff or equipment required for that step of the workflow. For example to start the procedure one needs to have the patient in the procedural room, he/she should be under anesthesia, the medical team should be ready, and the correct equipment, scope for endoscopy for example should be plug in. An Agent-Based Model (ABM) implementation [1] [5] [3] takes into account all these constraints if any and manages the spatio-temporal behavior of agents, i.e patient, staff, key pieces of equipment.

The task is a process with a duration that needs to be described. One uses a variety of models such as a probabilistic one that gives the duration once and for all, or a dynamic process that depends on a number of external factors including team performance.

The probability model uses a probability distribution that could be modulated by patient comorbidity and team past performance.

A dynamic modeling process with an ABM may use a set of discrete differential equations as in [3].

The progression 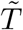 of the task *k* for the agent *i*, noted 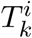, from 0 to 1 is described by an ordinary differential equation with the right hand side depending on the team skills. 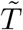 is set to 0 if the task is not completed, i.e. 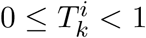, and 1 otherwise. M is a sparse matrix that corresponds to the directed graph of Figure 3. The master equation that provides the time evolution of the state of the graph of tasks 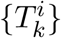 handled by the team *S*_*i*_ that advances the task *T*_*q*_ at time step *q* is:

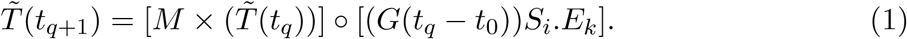

Here *×* denotes the sparse matrix vector product, and *°* the vector product component-wise, and. the product of a vector by a scalar.

The ABM model has three components:

- 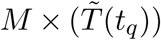 where *M* is a sparse matrix that expresses the dependency on previous tasks.
- *G*(*t*_*q*_ *− t*_0_)*S*_*i*_ reflects the time-dependent progression of the individual task.
- 0 ≤ *E*_*k*_ *≤* 1 is a positive factor representing a penalty for the environment conditions. It may represent the limitation resulting from shared equipment or specific overload of the GI lab due to epidemic or crisis.

In case of multiple paths coming out from a specific node in the workflow graph, as it would be the case for a GI lab in a Hospital environment, the path is activated according to a probability law or conditional probability similar with a Bayesian process.

##### Implementation of the Digital Twin

To go from the model to a digital twin one needs to fit the model to the observation in such a way that it can support heuristic computer reasoning. We will refer in the following to the overall architecture of Figure 4.

The underline model can be as simple as a Markovian process such as in [4] or an ABM as in [3] in this paper. Each probability law or discrete differential equation have generic construction relying on parameters. For example the shape of the distribution is given by its moments. The discrete differential equation (1) has coefficients.

Let us denote 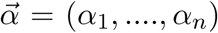 the n set of parameters defining the model. This set of parameter can include indicator of each provider and its team performances as well as duration of processes as a function of the distribution of staff in the peri-operative area. To build a digital twin of the workflow, the unknown vector parameter 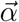 is obtained by fitting the model output to the observation; the fitting is solution of a (stochastic) minimization process as in [3, 7, 4]:

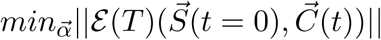

for a set of initial condition 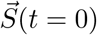, known control conditions 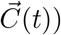 and duration *T*. In our notation the digital twin gives the simulation operator:

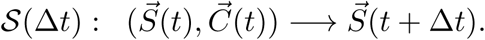

The predictive capability of this operator is essential to the success of computational heuristic reasoning; For robustness one has to:

- verify in real time that the digital twin runs properly and sends alert and/or makes correction automatically based on redundancy of input channel of information.
- double check the digital twin prediction off line to prevent errors due to delay in communication feed of input channels.
- verify and validate the prediction against observation by computing 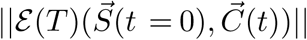.
- calibrate as needed the digital twin with the minimizing process as described above.
- exercise heuristic reasoning as described in the next section to support the decision of the manager:
  - in the short time scale (that requires restarting the digital twin in the middle of the day with hybrid inputs from EHR and sensor data)
  - in the long time scale that can start form a clean initial state before clinic start.

The digital twin uses a data base of observation with a large number of clinical days and time range that keeps track of all input channels. This set should be chosen in such a way that there is no under fitting, i.e the number of parameters to recover is much less than the number of conditions, and the workflow operates essentially with a stable infrastructure.

If the infrastructure changes a lot because of new providers coming in or providers leaving or new generation of equipment, etc… the digital twin needs to be fitted again. This can be done implicitly by monitoring if the prediction quality deteriorates with time.

In our method the digital twin is continuously fitted and the 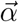 evolution can be interpreted as a learning process that catches changes in the infrastructure or evolution of the behavior of staff.

Decision are made through the change of the control variables 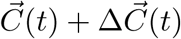.

Those changes should be bounded by ethical principle, i.e. admissible behavior, i.e admissible uses of the infrastructure

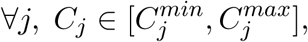

assuming continuity of error estimate with respect to control variable.

For example, there should be an upper limit on patient wait at each stage, or on overtime of staff.

*Note:* the *C*_*j*_ bounds may evolve as a function of the state variable that addresses staff/patient mental state or patient outcome, or specific environmental condition like a pandemic.

Let us described next how our mathematical framework makes heuristic reasoning that mimics *thought experiments* [2, 9] amenable to known optimization and data mining algorithm.

#### 2 Mathematical Formulation of *digital thought experiments* using the classification of Yeates’s 2004

##### Prefactua

”What will be the outcome if event X occurs?”

The abstract formulation of event X is a sudden change of the state variable 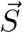, denoted 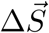 mathematical formation of this question is

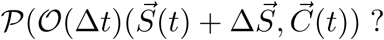

The algorithm to answer that problem is based on a forward digital twin run:

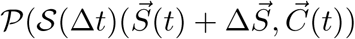

##### Counter Factual

”What might have happen if X had happened instead of Y?”

The mathematical formulation of this question is the same as above provided that 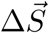 denotes the difference on the state variable between the sudden change of state variable switching from event X to event Y.

##### Semi-Factual

”Even though X occurs instead of Y would Z still occurs?”

Let’s assume that event Z corresponds to the *j* component of the projected value in the objective space. Using 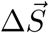 question defined as above, the mathematical formulation of this question would be:

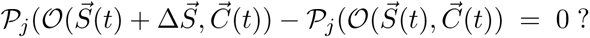

We answer this question by substituting the digital twin simulation to the observation.

##### Predictive

“Can we provide forecasting from stage Z?”

To formulate this question in a more rigorous way, we need to specify how further in time we like this prediction to be, i.e set Δ*t*, and how accurate should be this prediction to be valuable. Let’s assume 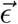 to be the tolerance for an admissible prediction value in the objective value space. The mathematical formulation of the question is:

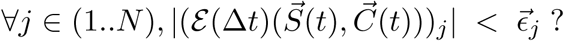

If this inequality is satisfied the prediction is correct for each component of the objective function. On one end, we check that inequality comparing observation with simulation and starting from some specific state value in the region of interest. On the other hand, we use the continuity of the error estimate with respect to state variable and this past observation to answer that question for any state values closer enough to *S*(*t*).

##### Hind Casting

”Can we provide forecasting from stage Z with new event X?”

This question is no different than the previous one except it applies to 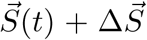 where 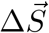 stands for new event X:

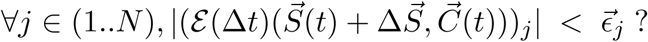

We use the same learning process from past experience described above to handle that problem.

##### Retrodiction

“past observations, events and data are used as evidence to infer the process(es) that produced them”

We start from a past observation:

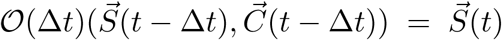

that has been tracking the state variable evolution in time for a given control variable. We verify that the model has been predictive:

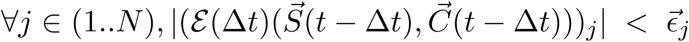

We assume that the error estimate is continuous with respect to the state variable.

The mathematical formulation of retrodiction can be done in many different ways depending on the level of causality we are looking for. In its simplest form, we look for the variable component that changes significantly the outcome, i.e.

Find *j ∈* (1..*N*), such that 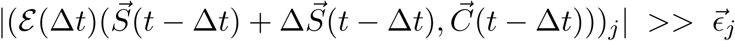 ?

This problem is amenable to standard optimization techniques.

A more sophisticated analysis would involve a non linear sensitivity analysis on all potential events or combination of events represented by 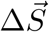 in a 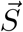, to be define.

##### Backcasting

”Moving backwards in time, step-by-step, in as many stages as are considered necessary, from the future to the present to reveal the mechanism through which that particular specified future could be attained from the present”

The mathematical formulation of that question can be derived from the above one; For example let’s assume that

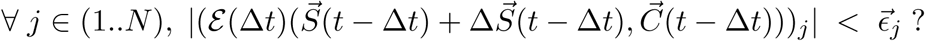

One can move one back step further to identify an event that would change the outcome:

Find *j ∈* (1..*N*), such that 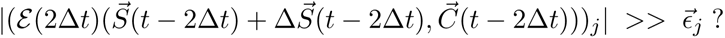 or repeat the process backward in time until such event exists. This would assume that the validity of the prediction holds for that many time steps backward, i.e.

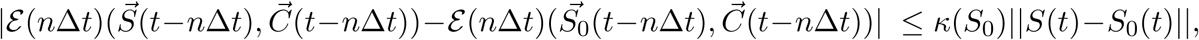

where n is the number of back steps Δ*t* involved and *S*_0_ is the state variable in the region of interest for the state of variable in the backward analysis.

## Notes

### Author Declarations

Ethics committee of the Office of Human Research at The George Washington University waived ethical approval for this work. They determined that: "Regarding the above referenced study, a determination has been made that this project does not meet the definition of human subjects research. That is, a living individual about whom and investigator 1) obtains data through intervention or interaction or 2) private identifiable information. This determination is being made because the study involves analyzing data to improve workflow efficiency in outpatient centers specialized in gastroenterology procedures. The study will not involve human subject interaction and/or any identifiable data.” The determination was made by Lacey Maddox, Research Compliance Associate at GWU.

